# Inequality in healthy lifespan following surgery: a longitudinal population study

**DOI:** 10.64898/2026.04.25.26351729

**Authors:** Yize I Wan, Rupert M Pearse, John R Prowle

**Affiliations:** William Harvey Research Institute, Queen Mary University of London, London, UK, EC1M 6BQ; Acute Critical Care Research Unit, Royal London Hospital, Barts Health NHS Trust, London, UK, E1 1FR

**Keywords:** surgery, long-term disease, socioeconomic deprivation, ethnicity, health inequality

## Abstract

**Background:** Surgery is a widely used treatment option but the impact of surgery on long-term disease across socioeconomic groups is unknown.

**Methods:** Longitudinal population study using linked primary and secondary care data describing adults (≥18 years) in England recorded in the Clinical Practice Research Datalink (CPRD) between 1st January 2012 and 31st December 2021. Socioeconomic deprivation was defined using the Index of Multiple Deprivation (IMD). The exposure was surgery and primary outcome was long-term disease. Data are presented as n (%), median (IQR), and adjusted hazards ratios (HR) with 95% confidence intervals.

**Findings:** Of 18,329,659 people, 8,951,145 (48·8%) underwent surgery. 78·6% of index surgeries were elective (n=7,032,475), 21·4% were emergency (n=1,918,670). Amongst surgical patients, 4,741,188 (52·0%) were women, 3,540,136 (39·6%) from the most deprived deciles (IMD 1-4) and 994,595 (11·1%) from a minority ethnic group. Age-standardised rates of surgery were higher in deprived individuals (comparative rate ratio IMD 1 vs. IMD 10 elective: 1·11 (95% CI 1·11-1·11), emergency: 1·54 (1·54-1·54)). Age at first surgery was 42 (27-60) years for elective and 42 (25-65) years for emergency surgery overall, but lower for people from IMD 1-4 (elective: 39 (26-57) years, emergency: 38 (24-60) years). Rates of long-term disease increased following both elective (baseline 19·6%, three years 24·5%) and emergency surgery (baseline 10·3%, three years 12·3%). Risk of new long-term disease following surgery increased with increasing levels of deprivation (IMD 1 vs. IMD 10 elective: HR 1·46 (1·45-1·48), emergency: HR 1·46 (1·44-1·48)).

**Interpretation:** Surgical treatment is strongly associated with the onset of long-term disease and factors which limit healthy life expectancy. Surgery occurs at a younger age among socioeconomically deprived groups and may be linked to health inequalities. Similar but more complex patterns of inequality were seen in minority ethnic groups.

**Funding:** Barts Charity and UK Academy of Medical Sciences.

**Research in Context:** *Evidence before this study:* The impact of surgery on long-term health outcomes beyond mortality and surgical complications such as persistent pain is unknown. People from deprived socioeconomic and minority ethnic groups experience increased risks of postoperative complications, readmissions, and death. We searched PubMed, for English language publications in adults aged over 19 years, over the last 10 years to 10th February 2026 using the following search terms: (surg* OR operat*) AND (long-term outcome OR chronic disease OR comorbidit* OR co-morbidit* OR multimorbidit* OR multi-morbidity*) AND (ethnic* OR race OR racial OR socio* OR depriv* OR ineq* OR disparit*). We identified 7,979 reports. To our knowledge, no previous studies have examined the development of long-term disease following surgery or differences in long-term outcomes following surgery between different socioeconomic and ethnic groups.

*Added value of this study:* This large national cohort study is to our knowledge the first to examine the relationship between surgery and onset of long-term disease. We included over 18·3 million individuals, of whom 48·8% underwent a surgical procedure during the study period. 5% of elective surgical patients and 2% of emergency surgical patients developed new long-term disease within three years of surgery. Accounting for differences in age, people from the most deprived decile experienced 11% higher rates of elective and 54% higher rates of emergency surgery compared to the least deprived. People in the most deprived decile underwent their first surgery nearly ten years earlier than those in the least deprived decile with a 46% higher risk of developing new long-term disease within three years of surgery. Although age standardised rates of surgery were lower, people from minority ethnic backgrounds underwent surgery for the first time up to sixteen years earlier that those from white backgrounds and had similarly had higher risk of developing new long-term disease at three years.

*Implications of all the available evidence:* Surgical treatments are associated with onset of long-term disease. This unexpected consequence should be considered when managing patients’ expectations when planning surgery with them. There are important inequalities in rates of both elective and emergency surgery, and the age this first occurs, between different socioeconomic and ethnic groups. Those from socioeconomically deprived backgrounds and minority ethnic groups undergo surgery at a younger age and are also at greater risk of developing long-term disease and hence reduced healthy life expectancy. One explanation for this may be differences in lifestyle and disease prevention behaviour. Surgery is therefore an important marker for inequalities in healthy life expectancy. The perioperative period is a key opportunity to better manage long-term health to reduce further inequalities. The patterns of these relationships are complex, and a more detailed understanding is needed to ensure that surgery can be better utilised as an opportunity to improve societal health.

## Introduction

Over 5·1 million NHS surgical procedures take place each year in the United Kingdom (UK).^1^ Surgery is seen as definitive and often curative treatment for a range of conditions, offering to improve either quantity of life, quality of life, or both. Around one in five in-patient surgical procedures will result in postoperative complications.^2^ Important complications such as infection, acute kidney injury and myocardial injury are associated with poor long term outcomes in the months and years which follow surgery.^3–5^ It is therefore plausible that surgery may be associated with the onset of long-term disease. However, there is little known about the impact of surgery on long-term health outcomes beyond mortality and specific complications.

People from deprived socioeconomic and minority ethnic groups experience worse outcomes following both elective and emergency surgery.^6–8^ Amongst surgical patients as for the general population, rates of pre-existing long-term disease increase with increasing deprivation and prevalence of long-term diseases vary between ethnic groups.^6–8^ Studies have shown that more deprived and minority ethnic groups often have increased surgical need but reduced access to elective procedures.^9^ Ethnicity has been recognised as a key risk factor in delayed diagnosis for several cancers, often resulting in surgery with increased complexity.^10^ Health inequalities in surgical care and outcomes may extend beyond the perioperative period and contribute to inequalities in long-term health.

Approximately 60% of the UK population will undergo a surgical procedure during their lifetime.^11^ The potential associations of surgery with long-term disease are not well understood. It is possible that the patterns of long-term disease after surgery represent an important example of health inequalities. We conducted an epidemiological study using linked routine primary and secondary care data in England to examine the onset of long-term disease following surgery and describe any differences between socioeconomic and ethnic groups.

## Methods

### Data source

We followed REporting of studies Conducted using Observational Routinely-collected Data (RECORD) guidelines (supplement). We used anonymised, routinely collected electronic health record data from the Clinical Practice Research Datalink (CPRD) database and linked datasets covering around 25% of the UK general population representative in terms of age, sex, and ethnicity.^12^ We included data from both CPRD GOLD and CPRD Aurum containing primary data collected using Vision® or EMIS® software systems respectively. Linked data to National Health Service hospitals in England was available from Hospital Episode Statistics (HES) up to 31st March 2021 including information on emergency department attendances using HES Accident and Emergency (HES A&E), hospitalisations using HES Admitted Patient Care (HES APC), and outpatient care (HES OP).^13^ Linked data to HES maternity were used to identify and exclude hospitalisation related to maternity care. Patient postcode linked socioeconomic deprivation measures were mapped using the composite 2019 English Index of Multiple Deprivation (IMD). Linkage to death registration data from the Office for National Statistics (ONS) was obtained. Postcode linked IMD was missing for 0·1% of the cohort. Linkage to all other datasets were complete. Access to data was approved via the CPRD research data governance process and this study was conducted in accordance with a pre-specified protocol (CPRD study protocol: 21_000720).

### Study cohort

We included all adults (≥18 years) residing in England registered with a primary care general practice (GP) between 1st January 2012 and 31st December 2022 who have not opted out of inclusion into CPRD and linked data sources. All individuals had at least 365 days of follow-up from date of GP registration and were followed up until the earliest of date of last data collection, transfer out of practice, or death.

### Definitions of key variables

The exposure was hospital admission for a surgical procedure during the follow-up period. Prior admissions outside of this window were not considered due to incomplete historical data recording of hospitalisation. Elective admissions were defined as waiting list, booked, and planned admissions. All other admissions were classed as emergency. Surgical procedures were identified and categorised by Office for Population Censuses Surveys Classification of Interventions and Procedures (OPCS) version 4·7 codes including procedures typically performed in an operating theatre under regional or general anaesthesia.^14^ Surgery related to obstetrics were excluded. The first surgical admission was identified as the index surgical admission. Individual-level relative measures of socioeconomic deprivation were assessed using IMD deciles (1st, most deprived). People with missing IMD were excluded from comparative analyses. Ethnicity was defined using the CPRD Ethnicity Record dataset (Asian, Black, Mixed/multiple, White, Other, Unknown) corresponding to UK Census 2011 higher-level categorisations.^15^ People with unknown ethnicity were included in comparative analyses as misclassification of non-White individuals into unknown or other ethnic groups occurs more frequently in emergency admissions. We examined age, sex, smoking history, alcohol intake, and obesity as additional baseline variables.

### Outcome measures

The outcome was the onset of new long-term disease at one and three years following surgery. We focused on long-term diseases included in the Charlson Comorbidity Index and previous multimorbidity studies.^16,17^ Diagnoses were identified from primary care records using phenotyping algorithms available online (full definitions in Table S1).

### Statistical analysis

We compared baseline characteristics between those who underwent elective and emergency surgery. We examined rates of surgery across the total CPRD cohort and age at first surgery based on IMD decile and ethnicity category. To account for potential differences in age structure between IMD deciles and between ethnic groups, we calculated comparative surgery ratios using age-standardised surgery rates based on ONS England population data for each year during the study period. We examined proportions of individuals who developed long-term disease over time including one year before surgery to three years after. Cox proportional hazards models censored for death were used to assess the relationship between IMD and ethnicity and development of long-term disease at one- and three years following surgery adjusted for age, sex, and baseline number of long-term diseases (categorised as none, one to two, more than two). Elective and emergency surgery were analysed separately and treated as time-dependent exposures. The analysis to three years excluded surgery from 1st January 2020 onwards. To account for non-linear effects of age, a restricted cubic spline model with five knots was used. People with unknown sex were excluded. Data are presented as mean (SD), median (IQR) or n (%). Effect measures are presented as hazards ratios (HR) with 95% confidence intervals (CI). All analyses were performed using RStudio 2025·05·1-513 and R version 4·5·1 (Rocky9) using OnDemand.^18^

### Sensitivity analyses

As cancer is an indication for surgery, we included a predefined sensitivity analysis excluding cancer as a long-term disease outcome.

## Results

Of a total 18,329,659 individuals within the CPRD cohort, 8,951,145 (48·8%) underwent a surgical procedure during the study period (Fig. S1). Baseline characteristics for the total cohort and stratified by the index surgical admission being elective or emergency are presented in Table 1. The majority of index surgeries were elective (n=7,032,475, 78·6%) compared to emergency (n=1,918,670, 21·4%). Numbers of included surgeries by category are listed in Table S2. People who underwent surgery for the first time as an emergency were older and a greater proportion were male. People who underwent first surgery electively were less likely to be current smokers, but a greater proportion had moderate to heavy alcohol use and obesity. Apart from cancer being more prevalent in people having first surgery electively, distribution of long-term diseases was similar between the two groups. Baseline characteristics stratified by elective or emergency surgery at any point are listed in Table S3. Compared to the elective surgery population, the emergency surgery population were younger, more likely to have history of smoking, alcohol intake and prevalence of most long-term diseases were higher. Cancer was more prevalent in people undergoing elective surgery.

**Table 1.**
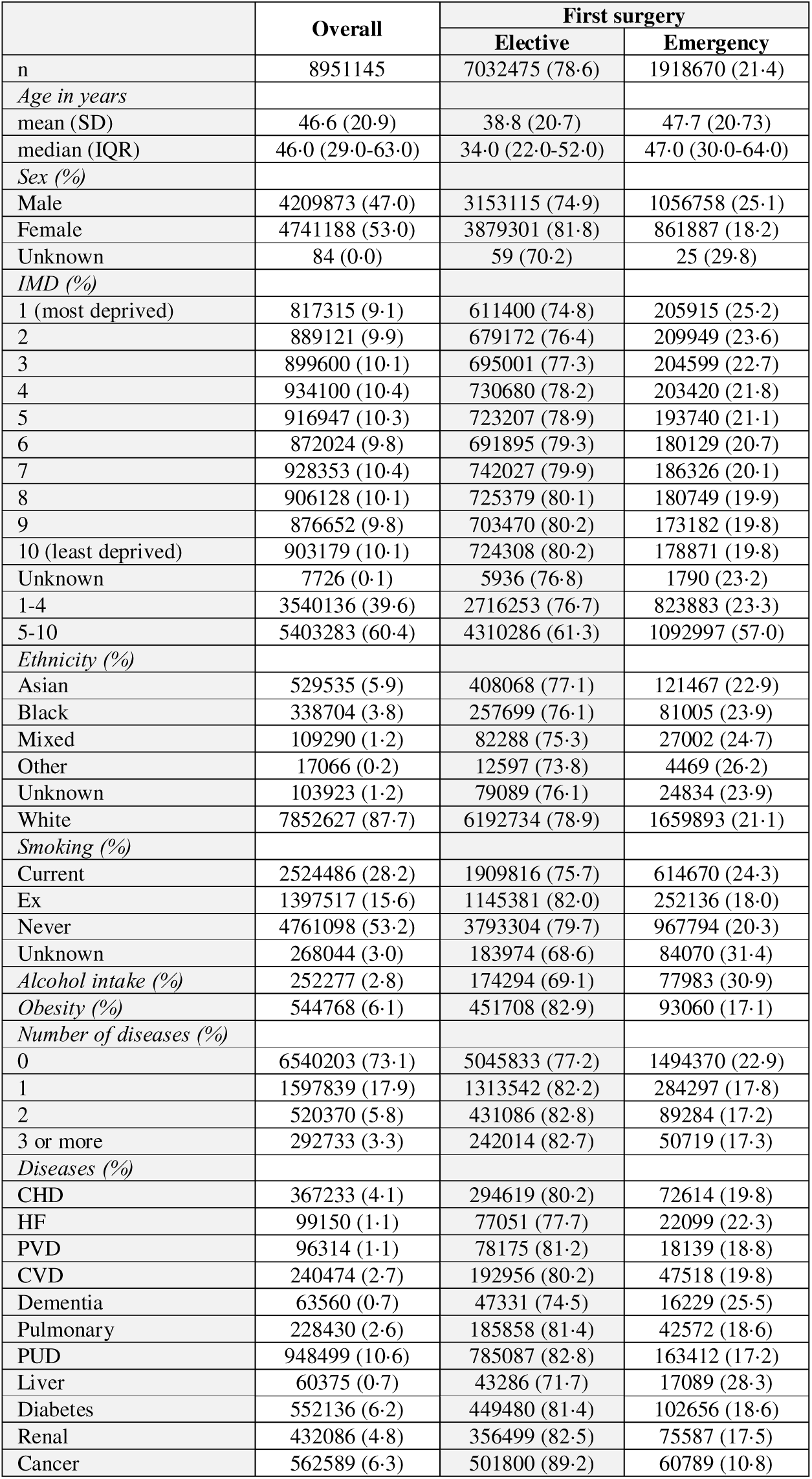
Baseline characteristics at start of follow-up on 01/01/2012, stratified according to first surgery being elective vs. emergency. All data presented as n (%) unless otherwise indicated, proportion by rows. IMD: index of multiple deprivation, CHD: coronary heart disease, HF: heart failure, PVD: peripheral vascular disease, PUD: peptic ulcer disease, CVD: cerebrovascular disease.

### IMD and rates of surgery and age at first surgery

In those undergoing surgery, 3,540,136 (39·6%) of people resided in the most deprived four IMD deciles 1-4 (Table 1). Absolute rates of surgery by age group within the total CPRD cohort did not change significantly over time with elective surgery being most frequent in those aged 70-89 years and emergency surgery being most frequent in those aged over 80 years (Fig. S2). Age-standardised surgery rates increased with increasing deprivation for both elective and emergency surgery but most markedly for emergency (Fig. S3). The average yearly comparative surgery ratio for IMD 1 compared to IMD 10 was 1·11 (95% CI 1·11-1·11) for elective and 1·54 (1·54-1·54) for emergency surgery (Fig. 1). Age at first surgery was 42 (27-60) years for elective and 42 (25-65) years for emergency overall, but lower in people from IMD 1-4 (elective: 39 (26-57) years, emergency: 38 (24-60) years) (Fig. 1 and Table S4). There was a difference in median age of 9 years between the most and least deprived deciles for elective (IMD 1: 38 (25-55) years, IMD 10: 47 (30-64) years) and 10 years for emergency surgery (IMD 1: 37 (25-58) years, IMD 10: 47 (28-69) years).

**Figure 1.**
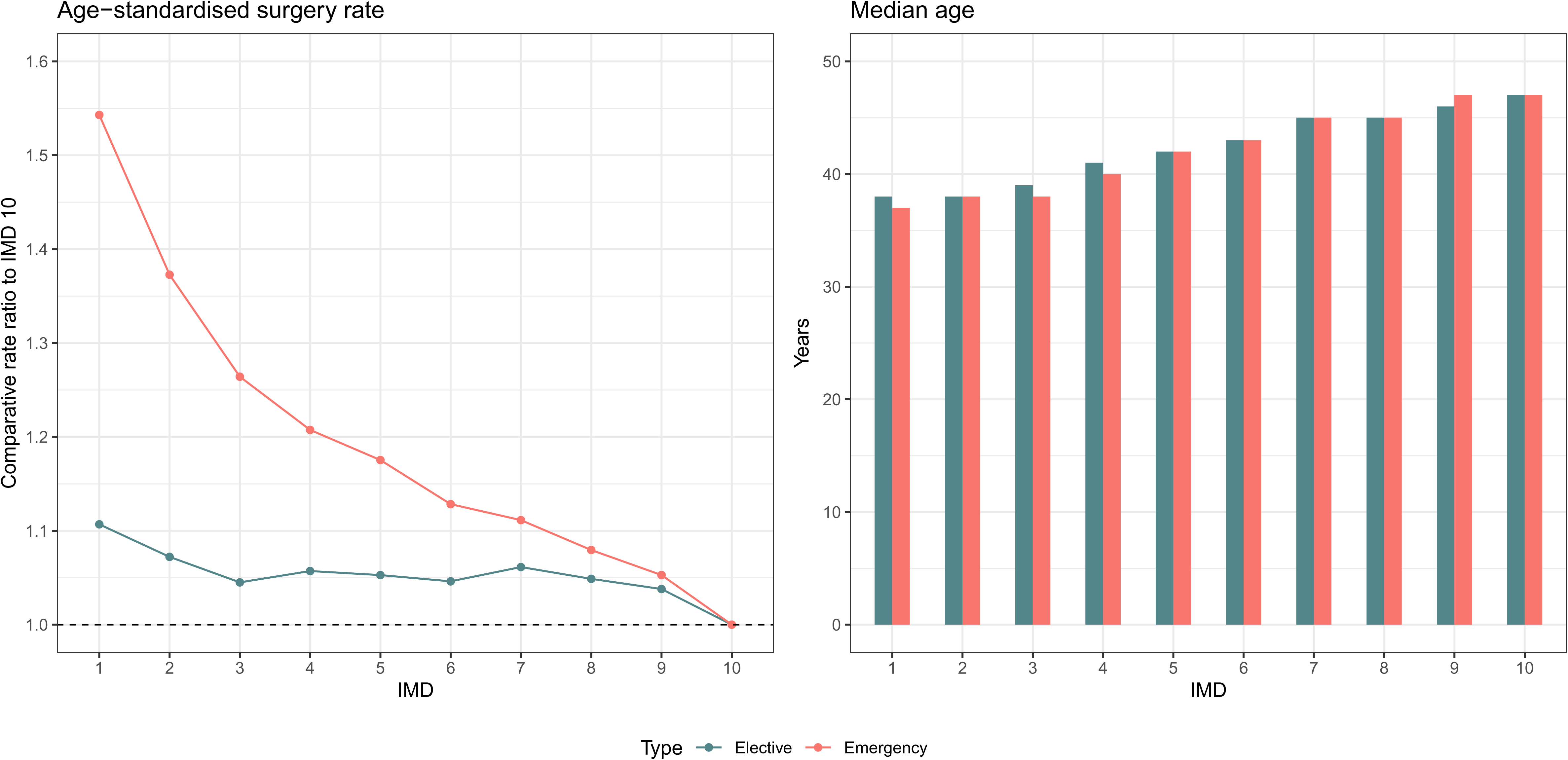
Comparative age-standardised surgery rate ratio and age at first surgical admission by IMD. Comparative ratio of the age-standardised surgery rate using ONS population data by type of surgery. Index of Multiple Deprivation (IMD) decile compared to IMD 10 (least deprived), 95% CI shown by error bars. Average yearly rate across the total study period.

### IMD and onset of long-term disease

Overall rates of baseline long-term disease at time of surgery were 19·6% for elective and 10·3% for emergency (Table S5). Baseline levels of long-term disease varied by level of deprivation for both types of surgery. Despite younger age, rates of long-term disease and multimorbidity were similar between individuals undergoing elective surgery in IMD 1 (19·6% and 5·7% respectively) and IMD 10 (20·4% and 5·1% respectively). For emergency surgery, rates of both long-term disease and multimorbidity were higher for more deprived patients (IMD 1: 11·5% and 4·8% vs. IMD 10: 9·9% and 4·0%). There was a greater increase in long-term disease following elective surgery compared to emergency at three years (5·0% elective, 2·1% emergency (Tables S5 and S6). Proportional increase in percentage of people with new long-term disease was higher in less deprived deciles following elective surgery (IMD 1: 24·8% vs. IMD 10: 25·2%) but higher in more deprived deciles following emergency surgery (IMD 1: 21·2% vs. IMD 10: 19·4%).

### IMD in Cox proportional hazards modelling

In surgical patients accounting for age, sex, and baseline number of long-term diseases, risk of new long-term disease following surgery increased with increasing levels of deprivation (Figs. 2 and S5). Compared to the least deprived (IMD 10), those residing in the most deprived decile undergoing elective surgery had 35% higher risk at one year (HR 1·35 (1·33-1·37)) and 46% higher risk at three years (HR 1·46 (1·45-1·48)) (Table S7). Following emergency surgery, effective sizes increased at one year: HR 1·41 (1·38-1·44) at one year, HR 1·46 (1·44-1·48) at three years (Table S8).

**Figure 2.**
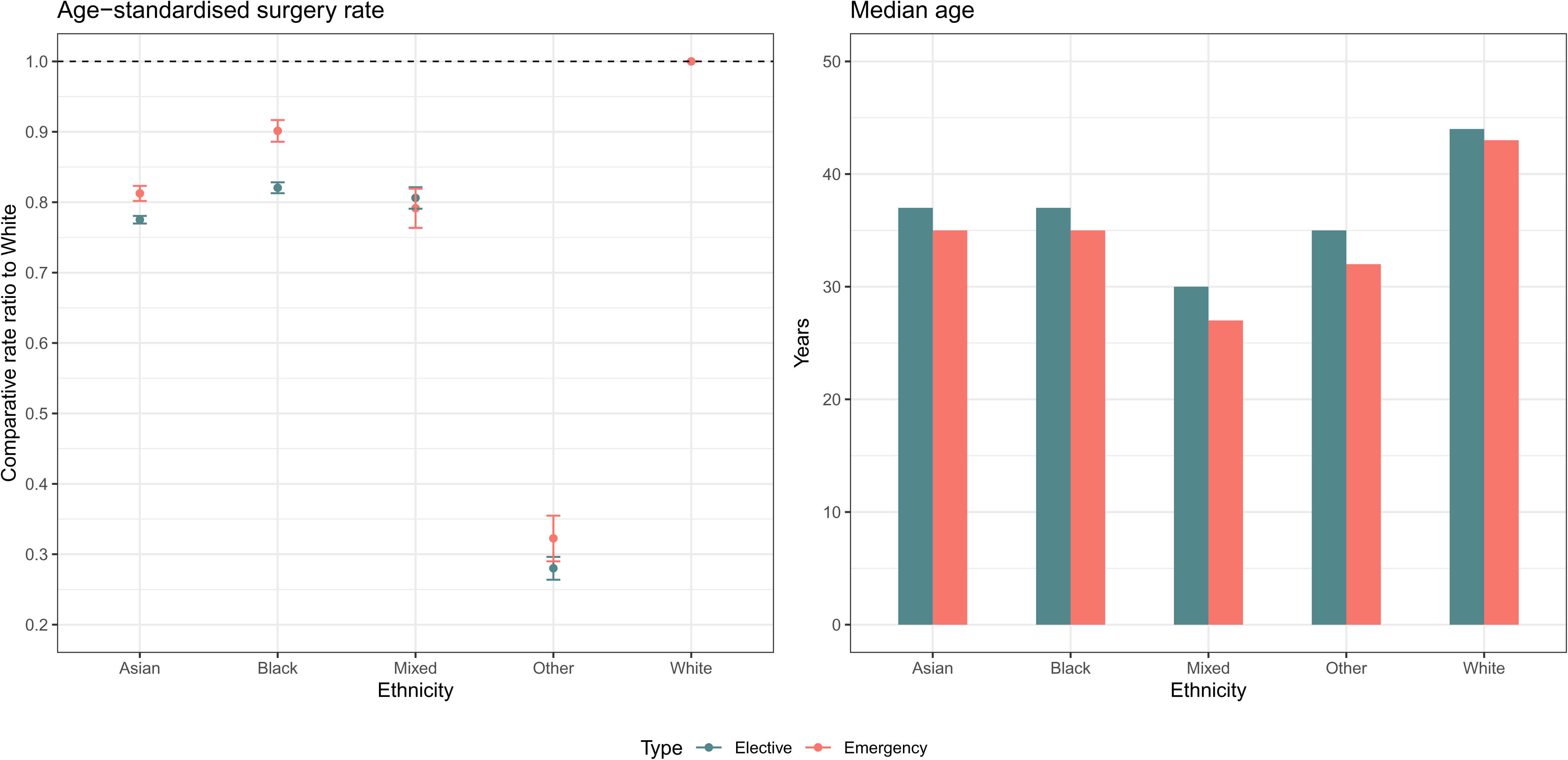
Comparative age-standardised surgery rate ratio and age at first surgical admission by ethnicity. Comparative ratio of the age-standardised surgery rate using ONS population data by type of surgery. Ethnicity compared to White, 95% CI shown by error bars. Average yearly rate across the total study period.

**Figure 3.**
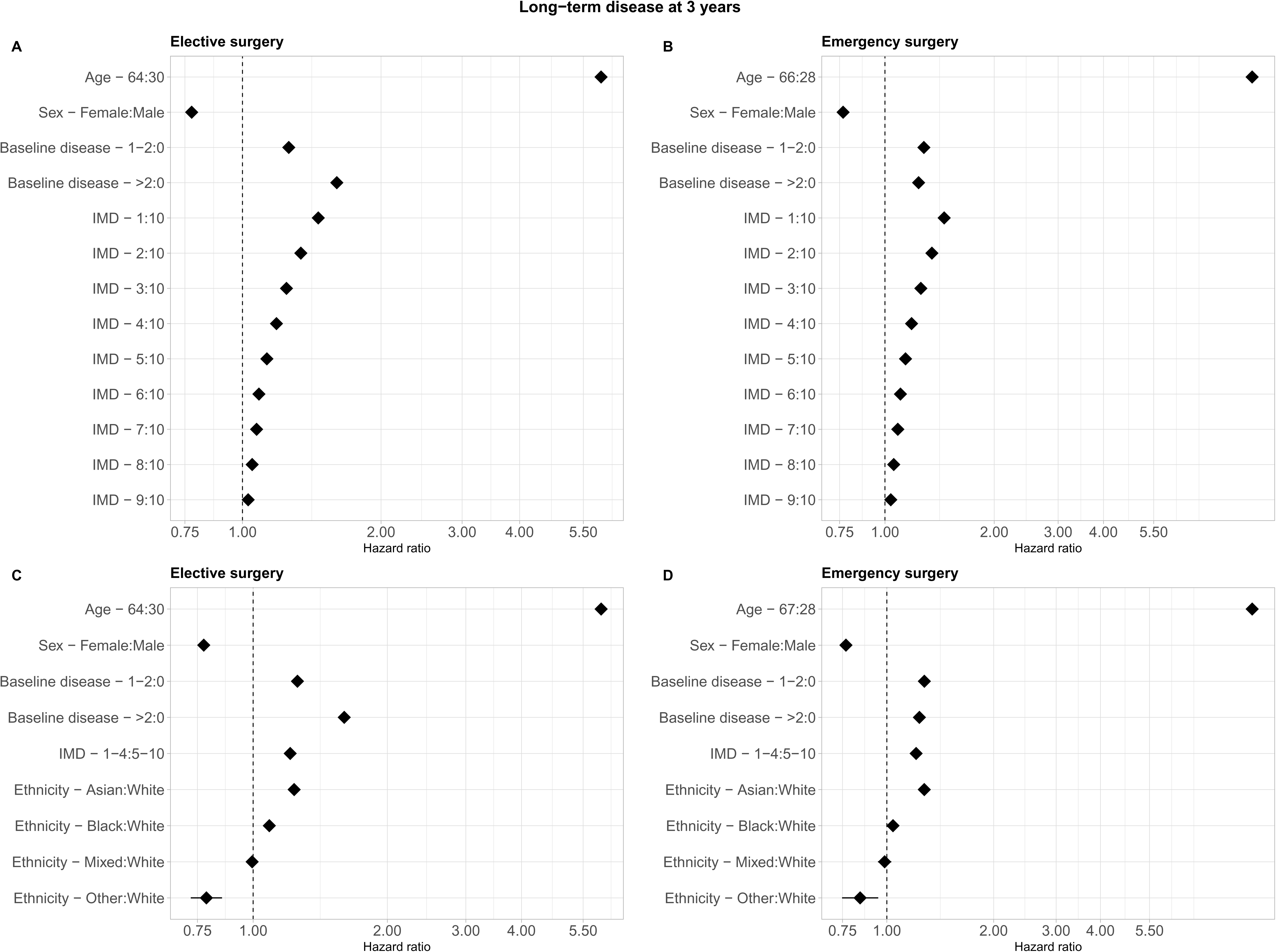
Forest plots showing adjusted HRs of developing long-term disease following surgery at three years. Panel A: Elective surgery by IMD decile, B: Emergency surgery by IMD decile, C: Elective surgery by ethnicity, D: Emergency surgery by ethnicity. Analysis of Index of Multiple Deprivation (IMD) decile compared to 10 (least deprived) included age, sex (excluding unknown), and baseline number of long-term diseases. Analysis of ethnicity compared to White included age, sex (excluding unknown), baseline number of long-term diseases, and most deprived IMD group (IMD 1-4). Hazard ratio for age comparing 75th to 25th centile. X-axis shown in log scale.

### Ethnicity and rates of surgery and age at first surgery

The ethnicity distribution was majority White (n=7,852,627, 87·7%), followed by Asian (n=529,535, 5·9%), Black (n=338,704, 3·8%), Mixed (n=109,290, 1·2%), and Other (n=17,066, 0·2%). 1·2% of were of unknown ethnicity (n=103,923) (Table 1). There were also consistent differences in age-standardised surgery rates across ethnic groups over time. Compared to White, rates were lower for people with all other ethnicities especially for elective surgery (Fig. S4). The average yearly comparative surgery ratios compared to White ethnicity were for Asian 0·78 (0·77-0·78) elective and 0·81 (0·80-0·82) emergency and for Black 0·82 (0·81-0·83) elective and 0·90 (0·89-0·92) emergency (Fig. 2). Age at first surgery was also lower in people from minority ethnic groups Patients from minority ethnic groups were significantly younger at first surgery compared to the White group with a difference in median age of 7-14 years for elective surgery and 8-16 years for emergency (elective: Asian 37 (26-52) years, Black 37 (26-50) years, White 44 (28-61) years; emergency: Asian 35 (25-52) years, Black 35 (24-48) years, White 43 (25-66) years) (Fig. 2 and Table S4).

### Ethnicity and onset of long-term disease

Baseline levels of long-term disease varied between ethnic groups for both types of surgery. White ethnicity patients, who were on average older than other ethnic groups, had the highest rates of long-term disease and multimorbidity for both elective and emergency surgery. Following elective surgery, people of Black ethnicity had the greatest proportional increase in new long-term disease (Black: 28·3%, White: 25·4%, Asian: 24·9%). Following emergency surgery, Black and Asian ethnic groups had greater proportional increase compared to White (Black: 25·2%, White: 19·8%, Asian: 22·9%).

### Ethnicity in Cox proportional hazards modelling

In surgical patients accounting for age, sex, baseline number of long-term diseases, and IMD, risk of new long-term disease following surgery increased with non-White ethnicity (Figs. 2 and S5). Patients of Asian and Black ethnicity had consistently higher risk of long-term disease compared to White patients for both types of surgery. For elective surgery, Asian patients had 1·14 (1·12-1·15) times greater risk at one year and 1·24 (1·22-1·25) times at three years, Black patients had 1·08 (1·05-1·10) times greater risk at one year and 1·09 (1·07-1·10) times at three years) (Table S7). Risk of long-term disease was highest for Asian patients following emergency surgery (HR 1·31 (1·29-1·34) at one year and 1·28 (1·26-1·29) at three years). Whereas for Black patients, risks were lower following emergency surgery compared to elective (HR 1·07 (1·05-1·10) at one year and 1·04 (1·02-1·06) at three years) (Table S8).

### Sensitivity analysis

Excluding cancer, increased rates of baseline long-term disease with increasing deprivation were also seen for those undergoing elective surgery (Table S5). At three years postoperatively, similar patterns in proportional increases in people with long-term disease were seen for elective surgery but not for emergency where increases in long-term disease were comparable between deprivation deciles (Table S6). Excluding cancer, Asian patients had the highest rates of baseline long-term disease and multimorbidity for elective surgery (Table S5).

Differences in proportional increases in people with long-term disease between ethnic groups remained in sensitivity analysis. Differences between IMD and ethnic groups for multimorbidity were even greater and remained in sensitivity analyses (Table S6). Risk of long-term disease in Cox modelling increased particularly for elective surgery such that patients in most deprived decile had 65% higher risk of long-term disease at one year and 67% at three years compared to the least deprived and Asian ethnicity patients had 45% higher risk at one year and 42% higher risk at three years compared to White (Tables S7 and S8, Figs. S6 and S7).

## Discussion

The principal finding of this longitudinal population study is that people from socioeconomically deprived and minority ethnic backgrounds experience a reduced healthy life expectancy associated with surgery. Our data does not demonstrate a causal link with surgery but given the known long-term health impacts of specific postoperative complications, it is likely that at least a proportion of this effect is due to the adverse effects of surgery. This observation is based on three linked findings from this analysis. Firstly, people from deprived IMD deciles experienced higher rates of surgery than expected for their age profile. Secondly, people living in the most deprived decile of the population undergoing first surgery are on average nine-to-ten years younger than those in the least. Thirdly, there is an increase in long-term disease following both elective and emergency surgery and the risk of developing new long-term disease at three years was 46% higher in the most compared to least deprived decile. Similarly, people from minority ethnic backgrounds undergoing surgery for the first time were on average sixteen years younger than those from white backgrounds and had higher risk of developing new long-term disease. However, those from minority ethnic backgrounds had lower age standardised rates of surgery, particularly elective, an observation that might reflect unequal use or access to health services. The underlying causes of these inequalities are unclear. However, it seems likely that differences in lifestyle and primary disease prevention in underserved groups may result in a greater, earlier need for surgical treatments in these population groups. These are to the best of our knowledge the first and largest published UK data to report on long-term health outcomes following surgery across socioeconomic and ethnic groups.

### Comparison to previous studies

Overall rates of surgery and proportions of elective compared to emergency surgery within our study are representative of the NHS surgical workload and similar to those previously reported.^11,19^ A few smaller UK observational cohort studies have examined differences in socioeconomic outcomes using data from surgical registries and national audits on surgical outcomes. These studies also found that increased deprivation was associated with increased rates of postoperative morbidity and mortality.^6,7,20^ Reported effect sizes vary but this is likely due to methodological differences in surgical urgency, surgical procedures included, risk adjustment methods, and outcome definitions. In these studies, patients in the most deprived quintiles also underwent surgery approximately ten years earlier than those in the least deprived and had a greater burden of long-term disease at baseline.^6,7^ However, overall ages at presentation were significantly younger in our study cohort by up to 20 years. This may be because we included an unselected population and examined rates of first surgery over a longitudinal period rather than incident cases at any time. There is also increasing evidence for ethnic disparities in surgery.^21–23^ However, these studies are limited by specific surgical disease indications, small sample sizes, and high rates of missing ethnicity records using single data sources. Findings from UK studies support socioeconomic and ethnic disparities reported by studies from other countries however comparison between countries is likely to be limited due to differences in baseline population demographics and healthcare systems resulting in differences in access to and delivery of surgery.

### Long-term disease following surgery

The intent for surgical treatment is almost exclusively to offer patients a greater quality of life and/or an extension in lifespan i.e. quantity of life. We found that even after the exclusion of cancer as a long-term disease due to cancer being a frequent indication for surgery, postoperative development of long-term disease still occurred at similar rates. People undergoing surgical procedures do not often regard their indication for surgery as a long-term health condition and consequently may not expect any change in their health status following surgery. Expectations of a rapid and uncomplicated recovery is particularly true for elective surgery.^24^ Patients with increased preoperative risk due to age, pre-existing long-term disease or multimorbidity and those undergoing higher risk procedures have been shown to account for disproportionate numbers of deaths after surgery and have increased healthcare use in the year following surgery.^25,26^ This is thought to be mediated through an increased risk of postoperative complications.^2^ However, even those without prior long-term diseases are at risk of postoperative complications, some of which have long-term health consequences.^5,27,28^ It may be that individuals have present for surgery with undiagnosed long-term diseases.^29^ This may be reflected in our finding that risk of new long-term disease was higher for elective compared to emergency surgery. The implications of surgery having a detrimental impact on long-term health and reducing health lifespan is important in being able to inform shared decision making and concerning for increased healthcare utilisation and subsequent burden on NHS services.

### Reasons for health inequalities

In the NHS, for the same disease presentation, patients across the UK are treated within comparable perioperative services, undergo the same type of surgical procedure, and receive similar levels of postoperative care. Despite this, we found evidence of socioeconomic and ethnic disparities in rates of surgery, age at first surgery, and postoperative outcomes. These disparities are likely to be driven at least in part by wider social determinants of health as well as inequalities in access to and delivery of services within sociodemographic groups and by geography.^9,30,31^ More deprived and minority ethnicity patients present with worse baseline health status and more advanced disease.^6,7,10^ This may increase vulnerability to the stress response to surgery and risk of postoperative complications. Higher levels of socioeconomic deprivation and minority ethnicity are also associated with lower rates or participation in cancer screening programmes, receipt of prehabilitation intervention.^32,33^ Understanding whether this is related to barriers to access and/or reduced engagement with health services and the reasons for these require analysis of provision and receipt of both preventative care and treatments, waiting list times for elective services, as well as qualitative analysis of experiences and attitudes of the population. These findings also highlight the need for ongoing public health and policy initiatives to both identify interventions to improve outcomes for all patients and ways to individualise care through specific targeted interventions both before and after surgery for disadvantaged groups.

### Strengths and limitations

This study has used a large, high quality national dataset including all regions of England and is broadly representative of the general population. Our cohort consisted of a non-selective sample of the general population at low risk of selection bias compared to other surgical cohorts. We followed a pre-specific statistical analysis plan and included both elective and emergency surgery over an 11-year period. We had near complete data linkage across primary and secondary care and to national statistics. However, there are some limitations. Socioeconomic deprivation has been assessed using a composite measure and therefore, we could not evaluate the direct effects of variations in other social determinants of health, differences in access to appropriate healthcare, or follow-up services. Area-level definitions of socioeconomic deprivation may underestimate the effect of inequalities compared to those observed at the individual level and do not account for duration of exposure to specific socioeconomic conditions. We were also not able to include individuals who were not registered with a GP or have a permanent address who are likely to have very high levels of deprivation. Like many datasets, ethnic categorisations were aggregated and do not reflect the considerable heterogeneity within ethnic groups. A small proportion of patients were classed as unknown or undisclosed ethnicity.

Misclassification of patients into both unknown or ‘other’ ethnic groups may occur more frequently in emergency consultations and short-term registrations where follow up data is incomplete leading to the observed markedly lower rates of surgery in these groups. While these data are presented for comparison, strong inferences about these ill-defined ethnic categories should not be drawn as assignment to these groups is likely to be biased toward patients with less clinical contact. There may also have be changes in IMD as people age and changes in how people identify by ethnic category over time which were not able to be assessed. The average age at first surgery reported in our analysis provides pragmatic value to clinicians and allows comparison between socioeconomic and ethnic groups in those undergoing surgery. However, this may not represent the age at which a person within the socioeconomic or ethnic group would on average first have surgery within the wider population. Reliance on clinical coding meant that assessment of important additional variables including occupation, lifestyle risk factors, variations in disease severity on admission, long-term disease management, and hospital process measures was not possible. Although obstetric related procedures were excluded, it is possible that some procedures related to reproductive medicine were included resulting in lower risk of outcomes in women of reproductive age. Finally, using observational data alone, we provide strong evidence for the association between but are not able prove a causal link between surgery and onset of long-term disease.

### Conclusions

Surgery is associated with an increased risk of developing long-term disease. There are important inequalities in this outcome, which are compounded by differences in rates of elective and emergency surgery, and the age this first occurs, between different socioeconomic and ethnic groups. Those from socioeconomically deprived backgrounds and minority ethnic groups undergoing surgery are younger when they first undergo surgery and are more likely to go on to develop long-term disease resulting in a short healthy life expectancy. This suggests that need for surgery may be an important marker for inequalities in healthy life expectancy and the perioperative period could be a key opportunity to better manage long-term health to reduce further inequalities. This has implications for shared decision making when planning surgery especially for elective surgery and for more disadvantaged population groups. Consideration of healthy life expectancy in the context of perioperative care may provide pathways for secondary prevention that have not previously been utilised and additional methods to improve societal health.

## Contributors

All authors were involved in study design, data application, and had access to all data in the study. YIW performed the statistical analysis, which was quality checked by RMP and JRP. All authors contributed to interpretation of the findings. YIW drafted the manuscript. All authors critically revised and approved the final manuscript for submission.

## Data sharing

All data supporting the findings in this study are provided by CPRD. Access to CPRD data is subject to approval by application via CPRD’s Research Data Governance process.

## Data statements

This research utilised Queen Mary’s Apocrita HPC facility, supported by QMUL Research-IT (doi:10.5281/zenodo.438045).

The CPRD Ethnicity Record sources underlying data from Hospital Episode Statistics (HES) Copyright © 2025, re-used with the permission of The Health & Social Care Information Centre. All rights reserved.

## Data Availability

All data supporting the findings in this study are provided by CPRD. Access to CPRD data is subject to approval by application via the CPRD Research Data Governance process.

## Acknowledgements

We acknowledge funding from Barts Charity (reference G-002354) and the Academy of Medical Sciences Starter Grant for Clinical Lecturers.

