## Supplemental file for "Inequality in healthy lifespan following surgery: a longitudinal population study"

### SUPPLEMENTARY FILE

Yize I Wan PhD,[1,2*] Rupert M Pearse MD,[1,2] John R Prowle MD,[1,2]

[1]William Harvey Research Institute, Queen Mary University of London, London, UK, EC1M 6BQ.
[2]Acute Critical Care Research Unit, Royal London Hospital, Barts Health NHS Trust, London, UK, E1 1FR.

*Correspondence to
Yize I Wan PhD,
Adult Critical Care Unit,
The Royal London Hospital,
Whitechapel
London E1 1FR


### Supplementary file

Cohort diagram

- Figure S1. Cohort definition.

Definitions of long-term disease

- Table S1. Definitions of long-term disease.

Definition of surgery

- Table S2. Types of surgery included.

Baseline characteristics by type of surgery for all surgeries

- Table S3. Baseline characteristics at start of follow-up for all elective and emergency surgery.

Age-standardised surgery rates

- Figure S2. Absolute surgery rate per 100,000 population per year.
- Figure S3. Age-standardised surgery rate by IMD decile.
- Figure S4. Age-standardised surgery rate by ethnicity.

Age at first surgery

- Table S4. Age at first surgery by IMD decile and ethnicity.

Development of long-term disease

- Table S5. Proportions with long-term disease and multimorbidity at baseline, one- and three years following elective and emergency surgery, by IMD decile and ethnicity.
- Table S6. Increase in long-term disease and multimorbidity at three years compared to baseline following elective and emergency surgery, by IMD decile and ethnicity.

Cox proportional hazards modelling

- Table S7. Association of IMD and ethnicity with development of long-term disease following elective surgery using Cox proportional hazards modelling.
- Table S8. Association of IMD and ethnicity with development of long-term disease following emergency surgery using Cox proportional hazards modelling.
- Figure S5. Forest plots showing adjusted HRs of developing long-term disease following elective and emergency surgery at one year.
- Figure S6. Forest plots showing adjusted HRs of developing long-term disease (excluding cancer) following elective and emergency surgery at one year.
- Figure S7. Forest plots showing adjusted HRs of developing long-term disease (excluding cancer) following elective and emergency surgery at three years.

RECORD checklist

**Figure S1. Cohort definition.** CPRD: clinical practice research datalink, HES: hospital episode statistics.

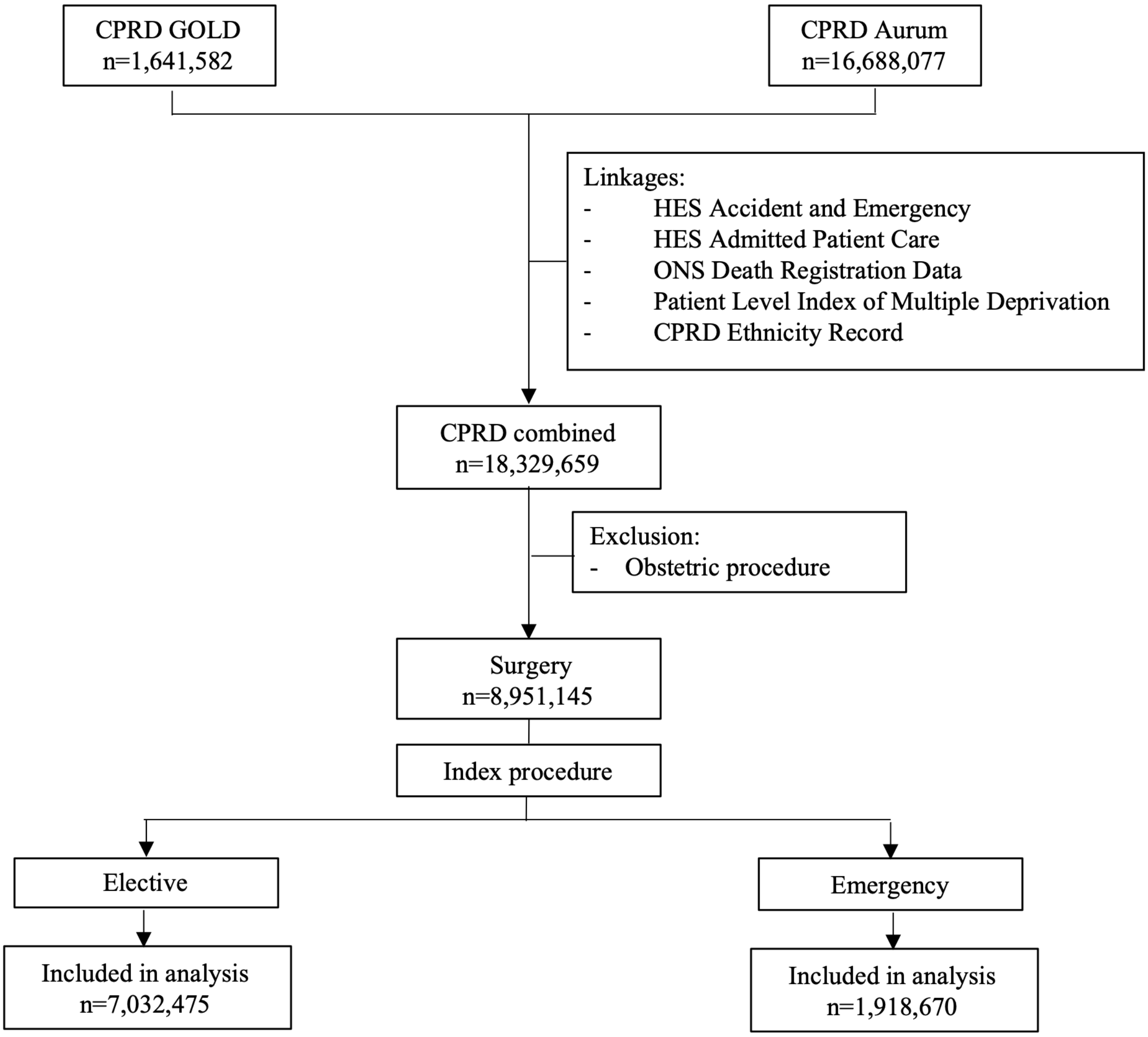

**Table S1. Definition of long-term disease and baseline risk factors.** CHD: coronary heart disease, HF: heart failure, PVD: peripheral vascular disease, PUD: peptic ulcer disease, CVD: cerebrovascular disease.

| **Disease category** | **Included conditions** |
|---|---|
| CHD | Coronary heart disease not otherwise specified |
| HF | Heart failure |
| PVD | Peripheral arterial disease |
| CVD | Ischaemic stroke, Stroke not otherwise specified, Transient ischaemic attack |
| Dementia | Dementia |
| Pulmonary | Bronchiectasis, COPD |
| PUD | Gastritis, Duodenitis, Gastro-oesophageal reflux disease, Peptic ulcer |
| Liver | Alcoholic liver disease, Chronic viral hepatitis, Fatty Liver, Hepatic failure |
| Diabetes | Diabetes |
| Renal | Chronic kidney disease, End stage renal disease |
| Cancer | Primary Malignancy: Biliary, Bladder, Bone, Bowel, Brain, Breast, Cervix, Multiple Sites, Oesophageal, Oropharyngeal, Kidney, Liver, Lung, Melanoma, Mesothelioma, Other, Ovary, Pancreas, Prostate, Skin, Stomach, Testis, Thyroid, Uterus, Adrenal. Secondary Malignancy: Bone, Bowel, Brain, Liver, Lung, Lymph Nodes, Peritoneum, Pleura |
| **Risk factor** | **Included conditions** |
| Smoking | Smoking |
| Alcohol | Alcohol misuse |
| Obesity | Obesity |

**Table S2. Types of surgery included.** All data presented as n (%), proportion by columns. GI: gastrointestinal, GU: genitourinary.

| | **All** | **Elective** | **Emergency** |
|---|---|---|---|
| Total | 8951145 | 7847419 | 3121732 |
| Bone | 592930 (6·6) | 300058 (3·8) | 562408 (18·0) |
| Breast | 197925 (2·2) | 202723 (2·6) | 15595 (0·5) |
| Cardiac | 248831 (2·8) | 169619 (2·2) | 229243 (7·3) |
| Cerebrovascular | 12647 (0·1) | 7459 (0·1) | 12668 (0·4) |
| Ear | 215770 (2·4) | 217061 (2·8) | 16145 (0·5) |
| Endocrine | 48084 (0·5) | 50551 (0·6) | 2891 (0·1) |
| Female lower GU | 142936 (1·6) | 132210 (1·7) | 30376 (1·0) |
| Female upper GU | 944189 (10·5) | 841078 (10·7) | 233089 (7·5) |
| HPB | 227049 (2·5) | 210213 (2·7) | 70178 (2·2) |
| Joint | 708411 (7·9) | 693185 (8·8) | 162191 (5·2) |
| Lower GI | 1047144 (11·7) | 767067 (9·8) | 547697 (17·5) |
| Major Vessel | 45462 (0·5) | 37021 (0·5) | 29966 (1·0) |
| Male GU | 335391 (3·7) | 322644 (4·1) | 47714 (1·5) |
| Muscle | 218459 (2·4) | 173212 (2·2) | 86510 (2·8) |
| Nasal | 245020 (2·7) | 243471 (3·1) | 34353 (1·1) |
| Neuro | 267508 (3·0) | 244342 (3·1) | 81820 (2·6) |
| Ocular | 726371 (8·1) | 773340 (9·9) | 61960 (2·0) |
| Oral | 578400 (6·5) | 577570 (7·4) | 65995 (2·1) |
| Orthopaedics | 33095 (0·4) | 35183 (0·4) | 4518 (0·1) |
| Other | 63649 (0·7) | 57867 (0·7) | 18721 (0·6) |
| Pharynx | 264356 (3·0) | 279226 (3·6) | 6840 (0·2) |
| Skin | 836356 (9·3) | 663675 (8·5) | 358075 (11·5) |
| Skull and Spine | 231510 (2·6) | 207858 (2·6) | 73327 (2·3) |
| Transplant | 1802 (0·0) | 1897 (0·0) | 80 (0·0) |
| Thoracic | 87741 (1·0) | 48672 (0·6) | 100553 (3·2) |
| Upper GI | 65155 (0·7) | 50220 (0·6) | 46737 (1·5) |
| Urological | 330271 (3·7) | 339180 (4·3) | 88081 (2·8) |
| Vascular | 234683 (2·6) | 200817 (2·6) | 134001 (4·3) |

**Table S3. Baseline characteristics at start of follow-up for all elective and emergency surgery.** All data presented as n (%) unless otherwise indicated, proportion by columns. IMD: index of multiple deprivation, CHD: coronary heart disease, HF: heart failure, PVD: peripheral vascular disease, PUD: peptic ulcer disease, CVD: cerebrovascular disease.

| | **Surgery** | |
|---|---|---|
| | **Elective** | **Emergency** |
| n | 7847419 | 3121732 |
| *Age in years* | | |
| mean (SD) | 47·7 (20·7) | 46·9 (22·3) |
| median (IQR) | 47·0 (30·0-64·0) | 45·0 (28·0-65·0) |
| *Sex (%)* | | |
| Male | 3587288 (45·7) | 1619905 (51·9) |
| Female | 4260063 (54·3) | 1501797 (48·1) |
| Unknown | 68 (0·0) | 30 (0·0) |
| *IMD (%)* | | |
| 1 (most deprived) | 698151 (8·9) | 326307 (10·5) |
| 2 | 767066 (9·8) | 332956 (10·7) |
| 3 | 780272 (10·0) | 324612 (10·4) |
| 4 | 816656 (10·4) | 328863 (10·5) |
| 5 | 806257 (10·3) | 317704 (10·2) |
| 6 | 769033 (9·8) | 296049 (9·5) |
| 7 | 822710 (10·5) | 310504 (10·0) |
| 8 | 802922 (10·2) | 300919 (9·6) |
| 9 | 777884 (9·9) | 287583 (9·2) |
| 10 (least deprived) | 799828 (10·2) | 293494 (9·4) |
| Unknown | 6640 (0·1) | 2741 (0·1) |
| 1-4 | 3062145 (39·1) | 1312738 (42·1) |
| 5-10 | 4778634 (60·9) | 1806253 (57·9) |
| *Ethnicity (%)* | | |
| Asian | 451343 (5·8) | 173957 (5·6) |
| Black | 287691 (3·7) | 116339 (3·7) |
| Mixed | 91760 (1·2) | 37032 (1·2) |
| Other | 13510 (0·2) | 5252 (0·2) |
| Unknown | 85002 (1·1) | 31530 (1·0) |
| White | 6918113 (88·2) | 2757622 (88·3) |
| *Smoking (%)* | | |
| Current | 2183204 (27·8) | 987130 (31·6) |
| Ex | 1269875 (16·2) | 479767 (15·4) |
| Never | 4187557 (53·4) | 1547454 (49·6) |
| Unknown | 206783 (2·6) | 107381 (3·4) |
| *Alcohol intake (%)* | 209932 (2·7) | 121657 (3·9) |
| *Obesity (%)* | 503886 (6·4) | 195384 (6·3) |
| *Number of diseases (%)* | | |
| 0 | 5624476 (71·7) | 2166939 (69·4) |
| 1 | 1463958 (18·7) | 575781 (18·4) |
| 2 | 483759 (6·2) | 227519 (7·3) |
| 3 or more | 275226 (3·5) | 151493 (4·9) |
| *Diseases (%)* | | |
| CHD | 339935 (4·3) | 182573 (5·8) |
| HF | 90599 (1·2) | 55460 (1·8) |
| PVD | 90524 (1·2) | 51999 (1·7) |
| CVD | 219196 (2·8) | 113530 (3·6) |
| Dementia | 52092 (0·7) | 33528 (1·1) |
| Pulmonary | 209718 (2·7) | 103687 (3·3) |
| PUD | 876072 (11·2) | 350008 (11·2) |
| Liver | 51596 (0·7) | 28811 (0·9) |
| Diabetes | 508748 (6·5) | 237885 (7·6) |
| Renal | 401328 (5·1) | 201093 (6·4) |
| Cancer | 544237 (6·9) | 202999 (6·5) |

**Figure S2. Absolute surgery rate per 100,000 population per year.** 2021 data to 31st March only.

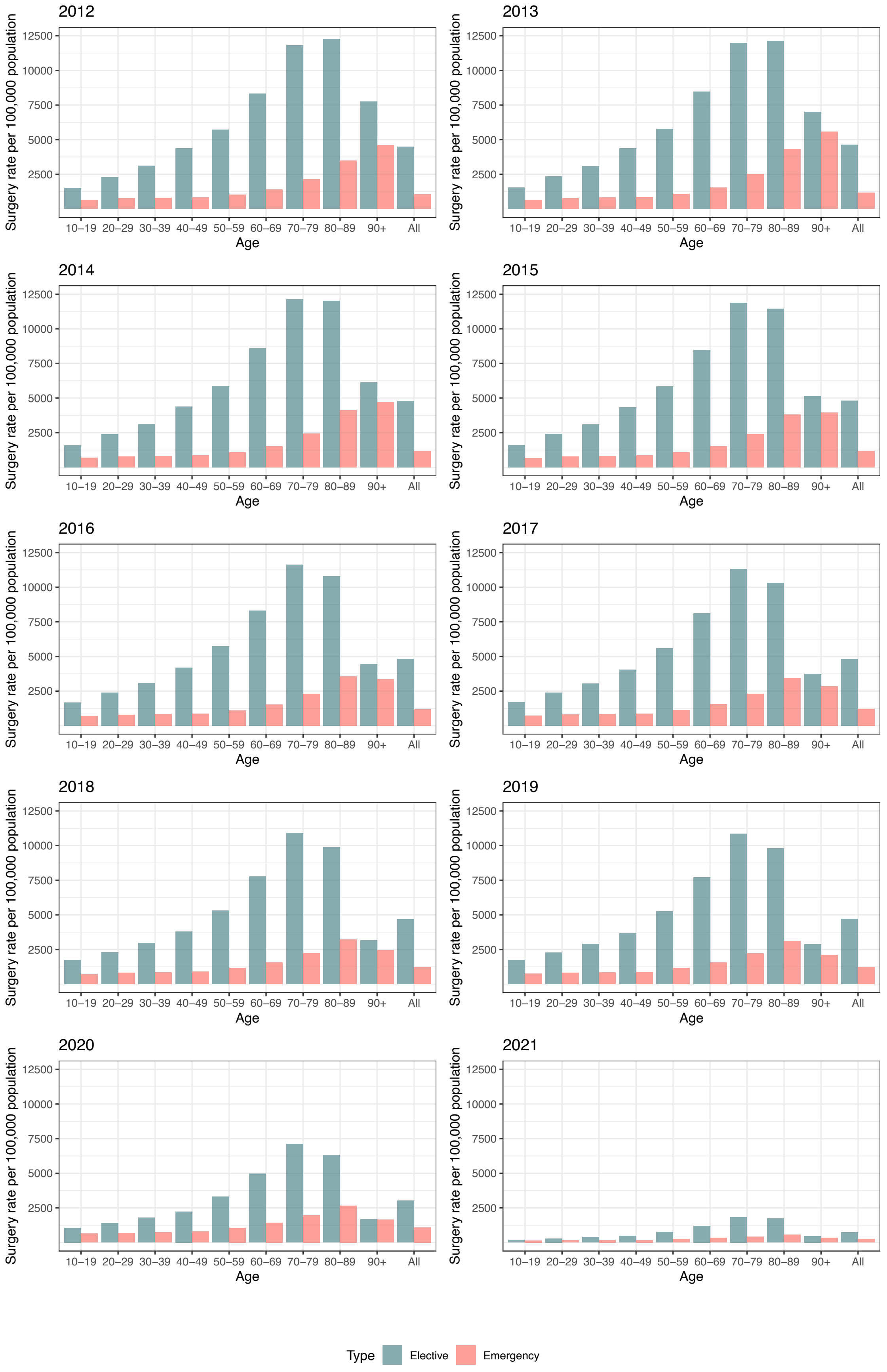

**Figure S3. Age-standardised surgery rate by IMD decile.** Comparative ratio of the age-standardised surgery rate using ONS population data by Index of Multiple Deprivation decile (IMD 1 most deprived), for each IMD decile compared to IMD 10 (least deprived), 95% CI shown by error bars. 2021 data to 31st March only.

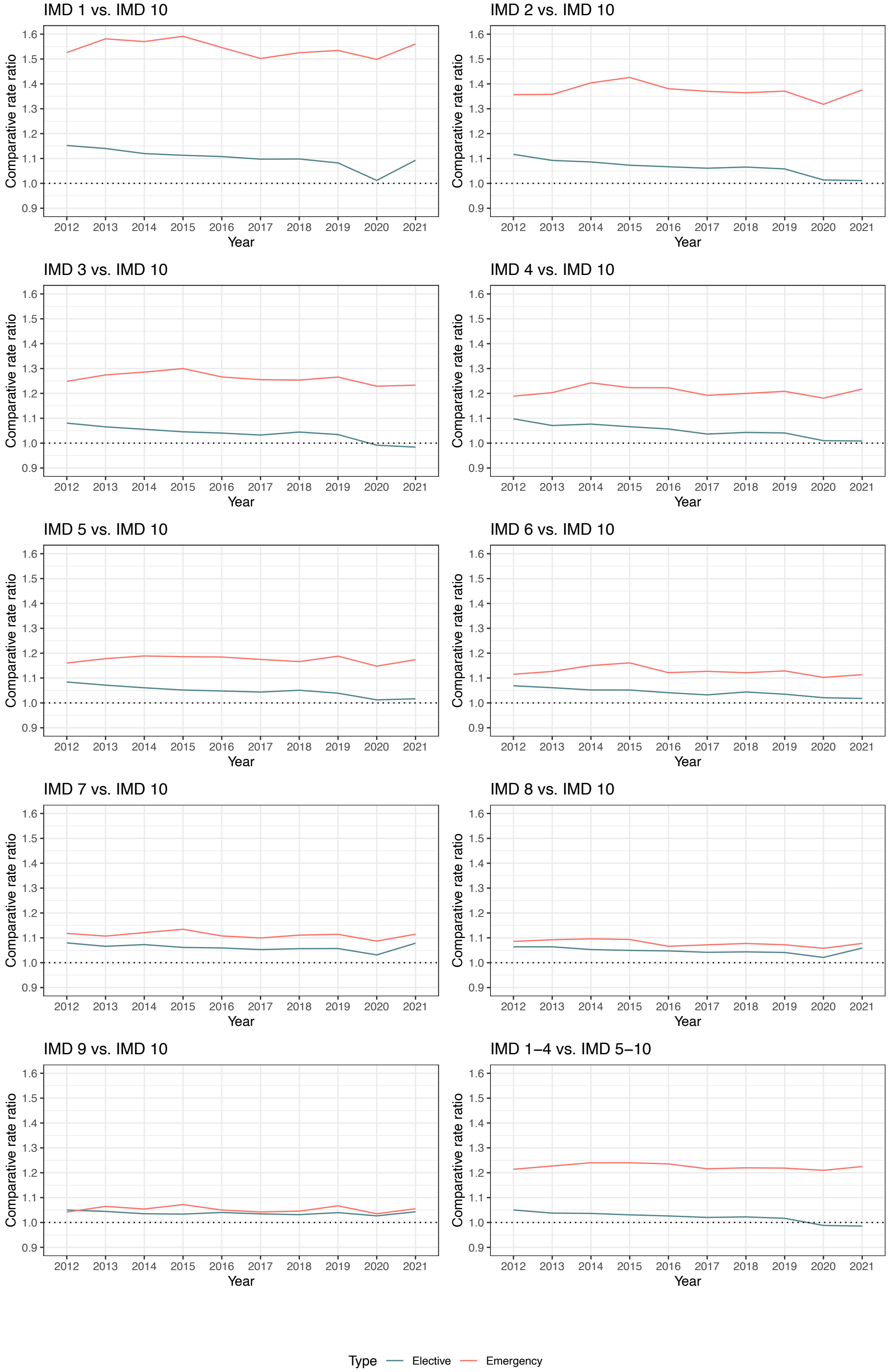

**Figure S4. Age-standardised surgery rate by ethnicity.** Comparative ratio of the age-standardised surgery rate using ONS population data by ethnicity, for each ethnicity compared to White, 95% CI shown by error bars. 2021 data to 31st March only.

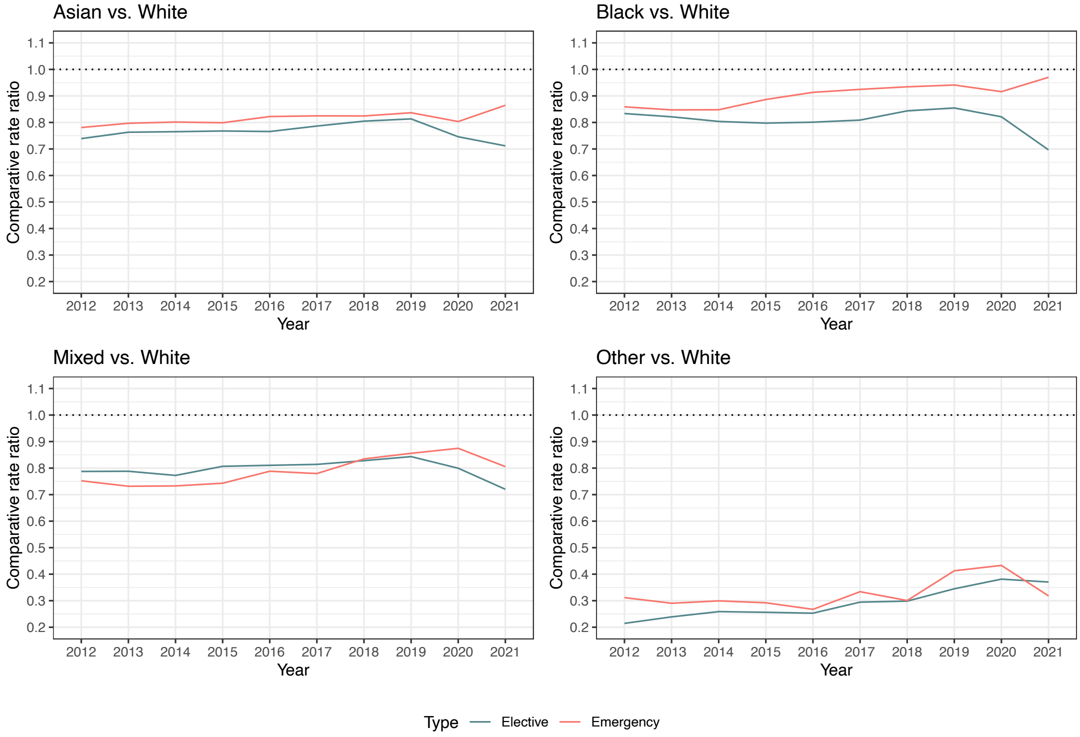

**Table S4. Age at first surgery by IMD decile and ethnicity.** Age in years presented as median (IQR). IMD: index of multiple deprivation.

| | Surgery | |
|---|---|---|
| | **Elective** | **Emergency** |
| All | 42 (27-60) | 42 (25-65) |
| *IMD* | | |
| 1 (most deprived) | 38 (25-55) | 37 (23-58) |
| 2 | 38 (25-56) | 38 (23-59) |
| 3 | 39 (26-57) | 38 (24-60) |
| 4 | 41 (27-59) | 40 (24-63) |
| 5 | 42 (27-60) | 42 (25-65) |
| 6 | 43 (28-61) | 43 (26-66) |
| 7 | 45 (29-62) | 45 (26-67) |
| 8 | 45 (29-62) | 45 (26-68) |
| 9 | 46 (29-63) | 47 (27-69) |
| 10 (least deprived) | 47 (30-64) | 47 (28-69) |
| 1-4 | 39 (26-57) | 38 (24-60) |
| 5-10 | 45 (29-62) | 45 (26-67) |
| *Ethnicity* | | |
| Asian | 37 (26-52) | 35 (25-52) |
| Black | 37 (26-50) | 35 (24-48) |
| Mixed | 30 (20-42) | 27 (18-39) |
| Other | 35 (25-47) | 32 (22-43) |
| Unknown | 38 (24-55) | 35 (20-57) |
| White | 44 (28-61) | 43 (25-66) |

**Table S5. Proportions with long-term disease and multimorbidity at baseline, one- and three years following elective and emergency surgery, by IMD decile and ethnicity.**

| | | Elective | | | | | | Emergency | | | | | |
|---|---|---|---|---|---|---|---|---|---|---|---|---|---|
| | | Long-term disease | | | Multimorbidity | | | Long-term disease | | | Multimorbidity | | |
| | | Baseline | 1 year | 3 years | Baseline | 1 year | 3 years | Baseline | 1 year | 3 years | Baseline | 1 year | 3 years |
| Main analysis | | | | | | | | | | | | | |
| **All** | | 19·1 | 21·8 | 24·5 | 5·1 | 6·2 | 7·8 | 10·3 | 11·4 | 12·3 | 4·2 | 5·0 | 5·7 |
| **IMD decile** | 1 (most deprived) | 19·6 | 21·6 | 24·4 | 5·7 | 6·8 | 8·5 | 11·5 | 12·8 | 13·9 | 4·8 | 5·7 | 6·5 |
| | 2 | 18·6 | 20·7 | 23·4 | 5·2 | 6·2 | 7·8 | 10·5 | 11·7 | 12·8 | 4·4 | 5·2 | 5·9 |
| | 3 | 18·6 | 20·7 | 23·4 | 5·0 | 6·0 | 7·5 | 10·1 | 11·2 | 12·2 | 4·1 | 4·9 | 5·6 |
| | 4 | 19·1 | 21·3 | 24·0 | 5·0 | 6·1 | 7·6 | 10·2 | 11·3 | 12·2 | 4·2 | 5·0 | 5·7 |
| | 5 | 19·4 | 21·7 | 24·4 | 5·1 | 6·1 | 7·7 | 10·2 | 11·3 | 12·2 | 4·2 | 5·0 | 5·7 |
| | 6 | 19·5 | 21·7 | 24·5 | 5·0 | 6·0 | 7·6 | 10·0 | 11·1 | 12·0 | 4·1 | 4·9 | 5·6 |
| | 7 | 20·1 | 22·5 | 25·3 | 5·2 | 6·3 | 8·0 | 10·2 | 11·3 | 12·2 | 4·2 | 5·0 | 5·7 |
| | 8 | 19·9 | 22·2 | 25·0 | 5·1 | 6·2 | 7·8 | 10·0 | 11·1 | 12·0 | 4·2 | 4·9 | 5·6 |
| | 9 | 20·3 | 22·7 | 25·5 | 5·1 | 6·2 | 7·8 | 10·1 | 11·2 | 12·1 | 4·2 | 4·9 | 5·6 |
| | 10 (least deprived) | 20·4 | 22·9 | 25·6 | 5·1 | 6·2 | 7·7 | 9·9 | 11·0 | 11·9 | 4·0 | 4·8 | 5·5 |
| **Ethnicity** | Asian | 19·1 | 21·0 | 23·8 | 5·0 | 5·9 | 7·3 | 8·7 | 9·8 | 10·7 | 3·4 | 4·1 | 4·7 |
| | Black | 15·6 | 17·5 | 20·0 | 3·5 | 4·4 | 5·5 | 7·2 | 8·2 | 9·0 | 2·5 | 3·0 | 3·5 |
| | Mixed | 10·5 | 11·8 | 13·5 | 1·9 | 2·3 | 3·0 | 4·7 | 5·4 | 5·9 | 1·2 | 1·5 | 1·8 |
| | Other | 9·5 | 10·8 | 12·4 | 1·7 | 2·1 | 2·4 | 3·6 | 4·4 | 4·9 | 0·9 | 1·1 | 1·3 |
| | White | 20·0 | 22·3 | 25·1 | 5·3 | 6·4 | 8·1 | 10·7 | 11·8 | 12·8 | 4·4 | 5·3 | 6·0 |
| Sensitivity analysis | | | | | | | | | | | | | |
| **All** | | 16·2 | 17·6 | 20·2 | 4·0 | 4·6 | 5·9 | 9·2 | 10·2 | 11·1 | 3·5 | 4·1 | 4·8 |
| **IMD decile** | 1 (most deprived) | 17·6 | 19·1 | 21·9 | 4·9 | 5·6 | 7·1 | 10·8 | 12·0 | 13·1 | 4·2 | 5·0 | 5·8 |
| | 2 | 16·5 | 17·9 | 20·6 | 4·3 | 5·0 | 6·3 | 9·8 | 10·8 | 11·9 | 3·8 | 4·5 | 5·2 |
| | 3 | 16·1 | 17·6 | 20·2 | 4·1 | 4·8 | 6·0 | 9·3 | 10·3 | 11·3 | 3·6 | 4·2 | 4·9 |
| | 4 | 16·1 | 17·6 | 20·2 | 4·0 | 4·7 | 5·9 | 9·2 | 10·2 | 11·1 | 3·5 | 4·2 | 4·8 |
| | 5 | 16·1 | 17·6 | 20·2 | 3·9 | 4·6 | 5·9 | 9·2 | 10·2 | 11·1 | 3·5 | 4·1 | 4·8 |
| | 6 | 15·8 | 17·2 | 19·8 | 3·8 | 4·4 | 5·6 | 8·9 | 9·9 | 10·8 | 3·3 | 4·0 | 4·6 |
| | 7 | 16·2 | 17·6 | 20·3 | 3·9 | 4·6 | 5·8 | 9·0 | 9·9 | 10·9 | 3·4 | 4·0 | 4·7 |
| | 8 | 15·8 | 17·3 | 19·8 | 3·8 | 4·4 | 5·6 | 8·8 | 9·7 | 10·6 | 3·4 | 4·0 | 4·6 |
| | 9 | 15·9 | 17·3 | 19·9 | 3·7 | 4·3 | 5·6 | 8·8 | 9·7 | 10·6 | 3·3 | 3·9 | 4·5 |
| | 10 (least deprived) | 15·6 | 16·9 | 19·5 | 3·6 | 4·2 | 5·3 | 8·5 | 9·4 | 10·3 | 3·1 | 3·7 | 4·3 |
| **Ethnicity** | Asian | 18·0 | 19·6 | 22·4 | 4·6 | 5·3 | 6·6 | 8·4 | 9·4 | 10·3 | 3·1 | 3·8 | 4·4 |
| | Black | 14·4 | 15·8 | 18·3 | 3·1 | 3·6 | 4·6 | 6·8 | 7·6 | 8·5 | 2·2 | 2·6 | 3·1 |
| | Mixed | 9·4 | 10·4 | 12·0 | 1·6 | 1·9 | 2·4 | 4·3 | 4·9 | 5·5 | 1·1 | 1·3 | 1·6 |
| | Other | 8·2 | 9·2 | 10·7 | 1·4 | 1·5 | 1·8 | 3·3 | 3·9 | 4·4 | 0·7 | 1·0 | 1·1 |
| | White | 16·3 | 17·7 | 20·4 | 4·1 | 4·7 | 6·0 | 9·5 | 10·5 | 11·5 | 3·6 | 4·3 | 5·0 |

**Table S6. Increase in long-term disease and multimorbidity at three years compared to baseline following elective and emergency surgery, by IMD decile and ethnicity.** Data showing proportional increase (absolute increase) from baseline in %. Proportional increase shown to allow comparison of IMD and ethnic groups with different baseline numbers of individuals.

| | | Elective | | Emergency | |
|---|---|---|---|---|---|
| | | **Long-term disease** | **Multimorbidity** | **Long-term disease** | **Multimorbidity** |
| Main analysis | | | | | |
| **All** | | 25·5 (5·0) | 51·6 (2·7) | 20·2 (2·1) | 35·7 (1·5) |
| **IMD decile** | 1 (most deprived) | 24·8 (4·9) | 48·7 (2·8) | 21·2 (2·4) | 36·0 (1·7) |
| | 2 | 25·7 (4·9) | 50·3 (2·6) | 21·4 (2·3) | 36·2 (1·6) |
| | 3 | 25·9 (4·9) | 50·5 (2·5) | 21·0 (2·1) | 35·8 (1·5) |
| | 4 | 25·8 (4·9) | 51·6 (2·6) | 20·1 (2·0) | 36·2 (1·5) |
| | 5 | 25·7 (5·0) | 52·4 (2·7) | 19·9 (2·0) | 35·2 (1·5) |
| | 6 | 25·8 (5·0) | 52·5 (2·6) | 20·2 (2·0) | 36·6 (1·5) |
| | 7 | 25·5 (5·1) | 52·1 (2·7) | 19·6 (2·0) | 35·9 (1·5) |
| | 8 | 25·5 (5·1) | 52·7 (2·7) | 19·6 (2·0) | 34·7 (1·4) |
| | 9 | 25·3 (5·1) | 52·8 (2·7) | 19·5 (2·0) | 35·0 (1·5) |
| | 10 (least deprived) | 25·2 (5·2) | 52·1 (2·6) | 19·4 (1·9) | 35·7 (1·4) |
| **Ethnicity** | Asian | 24·9 (4·8) | 47·2 (2·3) | 22·9 (2·0) | 39·6 (1·3) |
| | Black | 28·3 (4·4) | 55·1 (2·0) | 25·1 (1·8) | 39·4 (1·0) |
| | Mixed | 29·0 (3·0) | 58·4 (1·1) | 27·8 (1·3) | 45·0 (0·6) |
| | Other | 30·4 (2·9) | 41·1 (0·7) | 36·4 (1·3) | 50·7 (0·4) |
| | White | 25·4 (5·1) | 51·7 (2·7) | 19·8 (2·1) | 35·4 (1·6) |
| Sensitivity analysis | | | | | |
| **All** | | 25·1 (4·1) | 47·8 (1·9) | 21·2 (1·9) | 37·4 (1·3) |
| **IMD decile** | 1 (most deprived) | 24·4 (4·3) | 45·1 (2·2) | 21·4 (2·3) | 36·5 (1·5) |
| | 2 | 25·1 (4·1) | 46·4 (2·0) | 21·8 (2·1) | 37·0 (1·4) |
| | 3 | 25·3 (4·1) | 46·6 (1·9) | 21·4 (2·0) | 37·0 (1·3) |
| | 4 | 25·3 (4·1) | 47·4 (1·9) | 20·9 (1·9) | 37·6 (1·3) |
| | 5 | 25·3 (4·1) | 48·8 (1·9) | 20·7 (1·9) | 37·4 (1·3) |
| | 6 | 25·2 (4·0) | 49·0 (1·8) | 21·4 (1·9) | 38·5 (1·3) |
| | 7 | 25·1 (4·1) | 48·2 (1·9) | 21·0 (1·9) | 37·7 (1·3) |
| | 8 | 25·2 (4·0) | 49·1 (1·8) | 20·9 (1·8) | 36·8 (1·2) |
| | 9 | 25·0 (4·0) | 49·4 (1·8) | 21·1 (1·8) | 37·0 (1·2) |
| | 10 (least deprived) | 25·1 (3·9) | 49·1 (1·8) | 21·4 (1·8) | 38·8 (1·2) |
| **Ethnicity** | Asian | 24·4 (4·4) | 44·3 (2·0) | 22·9 (1·9) | 39·4 (1·2) |
| | Black | 26·6 (3·8) | 49·7 (1·5) | 25·0 (1·7) | 39·6 (0·9) |
| | Mixed | 28·3 (2·7) | 53·4 (0·8) | 27·3 (1·2) | 45·7 (0·5) |
| | Other | 30·4 (2·5) | 33·8 (0·5) | 35·6 (1·2) | 49·6 (0·4) |
| | White | 25·1 (4·1) | 48·0 (2·0) | 20·9 (2·0) | 37·1 (1·4) |

**Table S7. Association of IMD and ethnicity with development of long-term disease following elective surgery using Cox proportional hazards modelling.** Surgery as a time-dependent variable. Analysis to three years excluding surgery from 01/01/2020 onwards. *Variables included age, sex (excluding unknown), and baseline number of long-term diseases. [a]Variables included age, sex (excluding unknown), IMD most, and baseline number of long-term diseases.

| | | 1 year | | | 3 years | | |
|---|---|---|---|---|---|---|---|
| | | **Model** | **Hazards ratio (95% CI)** | **p value** | **Model** | **Hazards ratio (95% CI)** | **p value** |
| Main analysis | | | | | | | |
| **IMD decile** | 1 (most deprived) | Adjusted* (n=10413834, events= 334177) | 1·35 (1·33-1·37) | <0·0001 | Adjusted* (n=10032880, events= 733849) | 1·46 (1·45-1·48) | <0·0001 |
| | 2 | | 1·26 (1·24-1·28) | <0·0001 | | 1/34 (1·33-1·35) | <0·0001 |
| | 3 | | 1·17 (1·16-1·19) | <0·0001 | | 1·25 (1·23-1·26) | <0·0001 |
| | 4 | | 1·13 (1·11-1·15) | <0·0001 | | 1·19 (1·17-1·19) | <0·0001 |
| | 5 | | 1·08 (1·06-1·10) | <0·0001 | | 1·13 (1·12-1·14) | <0·0001 |
| | 6 | | 1·05 (1·04-1·07) | <0·0001 | | 1·09 (1·07-1·10) | <0·0001 |
| | 7 | | 1·04 (1·02-1·06) | <0·0001 | | 1·07 (1·06-1·08) | <0·0001 |
| | 8 | | 1·02 (1·01-1·04) | 0·002 | | 1·05 (1·04-1·06) | <0·0001 |
| | 9 | | 1·00 (0·99-1·02) | 0·97 | | 1·03 (1·02-1·04) | <0·0001 |
| | 10 (least deprived) | | Reference | - | | Reference | - |
| **Ethnicity** | Asian | Adjusted[a] (n=10296327, events= 331773) | 1·14 (1·12-1·15) | <0·0001 | Adjusted[a] (n=9922389, events= 729133) | 1·24 (1·22-1·25) | <0·0001 |
| | Black | | 1·08 (1·05-1·10) | <0·0001 | | 1·09 (1·07-1·10) | <0·0001 |
| | Mixed | | 0·99 (0·95-1·03) | 0·62 | | 0·99 (0·97-1·02) | 0·73 |
| | Other | | 0·82 (0·73-0·92) | 0·0006 | | 0·79 (0·72-0·85) | <0·0001 |
| | White | | Reference | - | | Reference | - |
| Sensitivity analysis | | | | | | | |
| **IMD decile** | 1 (most deprived) | Adjusted* (n=10413834, events= 209131) | 1·65 (1·62-1·68) | <0·0001 | Adjusted* (n=10032880, events= 566014) | 1·67 (1·65-1·69) | 0·0001 |
| | 2 | | 1·50 (1·47-1·52) | <0·0001 | | 1·50 (1·48-1·52) | 0·0001 |
| | 3 | | 1·37 (1·34-1·40) | <0·0001 | | 1·38 (1·36-1·40) | 0·0001 |
| | 4 | | 1·29 (1·26-1·31) | <0·0001 | | 1·29 (1·27-1·30) | 0·0001 |
| | 5 | | 1·20 (1·18-1·23) | <0·0001 | | 1·21 (1·20-1·23) | 0·0001 |
| | 6 | | 1·13 (1·11-1·15) | <0·0001 | | 1·14 (1·13-1·16) | 0·0001 |
| | 7 | | 1·11 (1·08-1·13) | <0·0001 | | 1·12 (1·10-1·13) | 0·0001 |
| | 8 | | 1·08 (1·06-1·10) | <0·0001 | | 1·09 (1·08-1·10) | 0·0001 |
| | 9 | | 1·03 (1·01-1·05) | 0·001 | | 1·05 91·04-1·07) | 0·0001 |
| | 10 (least deprived) | | Reference | - | | Reference | - |
| **Ethnicity** | Asian | Adjusted[a] (n=10296327, events= 207808) | 1·45 (1·42-1·47) | <0·0001 | Adjusted[a] (n=9922389, events= 562615) | 1·42 (1·40-1·44) | 0·0001 |
| | Black | | 1·19 (1·16-1·22) | <0·0001 | | 1·16 (1·14-1·18) | 0·0001 |
| | Mixed | | 1·10 (1·04-1·16) | 0·0003 | | 1·07 (1·03-1·11) | 0·0001 |
| | Other | | 0·97 (0·85-1·11) | 0·66 | | 0·86 (0·79-0·94) | 0·0006 |
| | White | | Reference | - | | Reference | - |

**Table S8. Association of IMD and ethnicity with development of long-term disease following emergency surgery using Cox proportional hazards modelling.** Surgery as a time-dependent variable. Analysis to three years excluding surgery from 01/01/2020 onwards. *Variables included age, sex (excluding unknown), and baseline number of long-term diseases. [a]Variables included age, sex (excluding unknown), IMD most, and baseline number of long-term diseases.

| | | 1 year | | | 3 years | | |
|---|---|---|---|---|---|---|---|
| | | **Model** | **Hazards ratio (95% CI)** | **p value** | **Model** | **Hazards ratio (95% CI)** | **p value** |
| Main analysis | | | | | | | |
| **IMD decile** | 1 (most deprived) | Adjusted* (n=4527307, events= 210789) | 1·41 (1·38-1·44) | <0·0001 | Adjusted* (n=4197316, events= 370360) | 1·46 (1·44-1·48) | <0·0001 |
| | 2 | | 1·34 (1·31-1·36) | <0·0001 | | 1·35 (1·33-1·37) | <0·0001 |
| | 3 | | 1·25 (1·22-1·27) | <0·0001 | | 1·26 (1·24-1·27) | <0·0001 |
| | 4 | | 1·17 (1·14-1·19) | <0·0001 | | 1·18 (1·17-1·20) | <0·0001 |
| | 5 | | 1·13 (1·11-1·16) | <0·0001 | | 1·14 (1·12-1·16) | <0·0001 |
| | 6 | | 1·11 (1·09-1·13) | <0·0001 | | 1·10 (1·09-1·12) | <0·0001 |
| | 7 | | 1·07 (1·05-1·09) | <0·0001 | | 1·09 (1·07-1·10) | <0·0001 |
| | 8 | | 1·04 (1·02-1·06) | <0·0001 | | 1·06 (1/04-1·07) | <0·0001 |
| | 9 | | 1·02 (1·00-1·04) | 0·04 | | 1·04 (1·02-1·05) | <0·0001 |
| | 10 (least deprived) | | Reference | - | | Reference | - |
| **Ethnicity** | Asian | Adjusted[a] (n=4482249, events= 209356) | 1·31 (1·29-1·34) | <0·0001 | Adjusted[a] (n=4156512, events= 368086 | 1·28 (1·26-1·29) | <0·0001 |
| | Black | | 1·07 (1·05-1·10) | <0·0001 | | 1·04 (1·02-1·06) | <0·0001 |
| | Mixed | | 1·03 (0·97-1·09) | 0·31 | | 0·99 (0·94-1·03) | 0·52 |
| | Other | | 1·01 (0·87-1·16) | 0·94 | | 0·84 (0·74-0·95) | 0·004 |
| | White | | Reference | - | | Reference | - |
| Sensitivity analysis | | | | | | | |
| **IMD decile** | 1 (most deprived) | Adjusted* (n=4527307, events= 175923) | 1·48 (1·45-1·51) | <0·0001 | Adjusted* (n=4197316, events= 322313) | 1·53 (1·51-1·56) | <0·0001 |
| | 2 | | 1·39 (1·36-1·42) | <0·0001 | | 1·40 (1·38-1·43) | <0·0001 |
| | 3 | | 1·28 (1·26-1·31) | <0·0001 | | 1·31 (1·28-1·33) | <0·0001 |
| | 4 | | 1·20 (1·17-1·22) | <0·0001 | | 1·22 (1·20-1·24) | <0·0001 |
| | 5 | | 1·15 (1·13-1·18) | <0·0001 | | 1·17 (1·15-1·19) | <0·0001 |
| | 6 | | 1·13 (1·11-1·16) | <0·0001 | | 1·13 (1·11-1·15) | <0·0001 |
| | 7 | | 1·08 (1·06-1·11) | <0·0001 | | 1·11 (1·09-1·12) | <0·0001 |
| | 8 | | 1·05 (1·03-1·07) | <0·0001 | | 1·07 (1·06-1·09) | <0·0001 |
| | 9 | | 1·03 (1·00-1·05) | 0·02 | | 1·05 (1·03-1·07) | <0·0001 |
| | 10 (least deprived) | | Reference | - | | Reference | - |
| **Ethnicity** | Asian | Adjusted[a] (n=4482249, events= 174823) | 1·42 (1·39-1·45) | <0·0001 | Adjusted[a] (n=4156512, events= 320421) | 1·36 (1·34-1·39) | <0·0001 |
| | Black | | 1·09 (1·06-1·12) | <0·0001 | | 1·07 (1·05-1·09) | <0·0001 |
| | Mixed | | 1·05 (0·99-1·12) | 0·13 | | 1·00 (0·96-1·05) | 1·00 |
| | Other | | 0·99 (0·85-1·16) | 0·91 | | 0·86 (0·76-0·98) | 0·02 |
| | White | | Reference | - | | Reference | - |

**Figure S5. Forest plots showing adjusted HRs of developing long-term disease following surgery at one year. Panel A: Elective surgery by IMD decile, B: Emergency surgery by IMD decile, C: Elective surgery by ethnicity, D: Emergency surgery by ethnicity.** Analysis of Index of Multiple Deprivation (IMD) decile compared to 10 (least deprived) included age, sex (excluding unknown), and baseline number of long-term diseases. Analysis of ethnicity compared to White included age, sex (excluding unknown), baseline number of long-term diseases, and most deprived IMD group (IMD 1-4). Hazard ratio for age comparing 75th to 25th centile. X-axis shown in log scale.

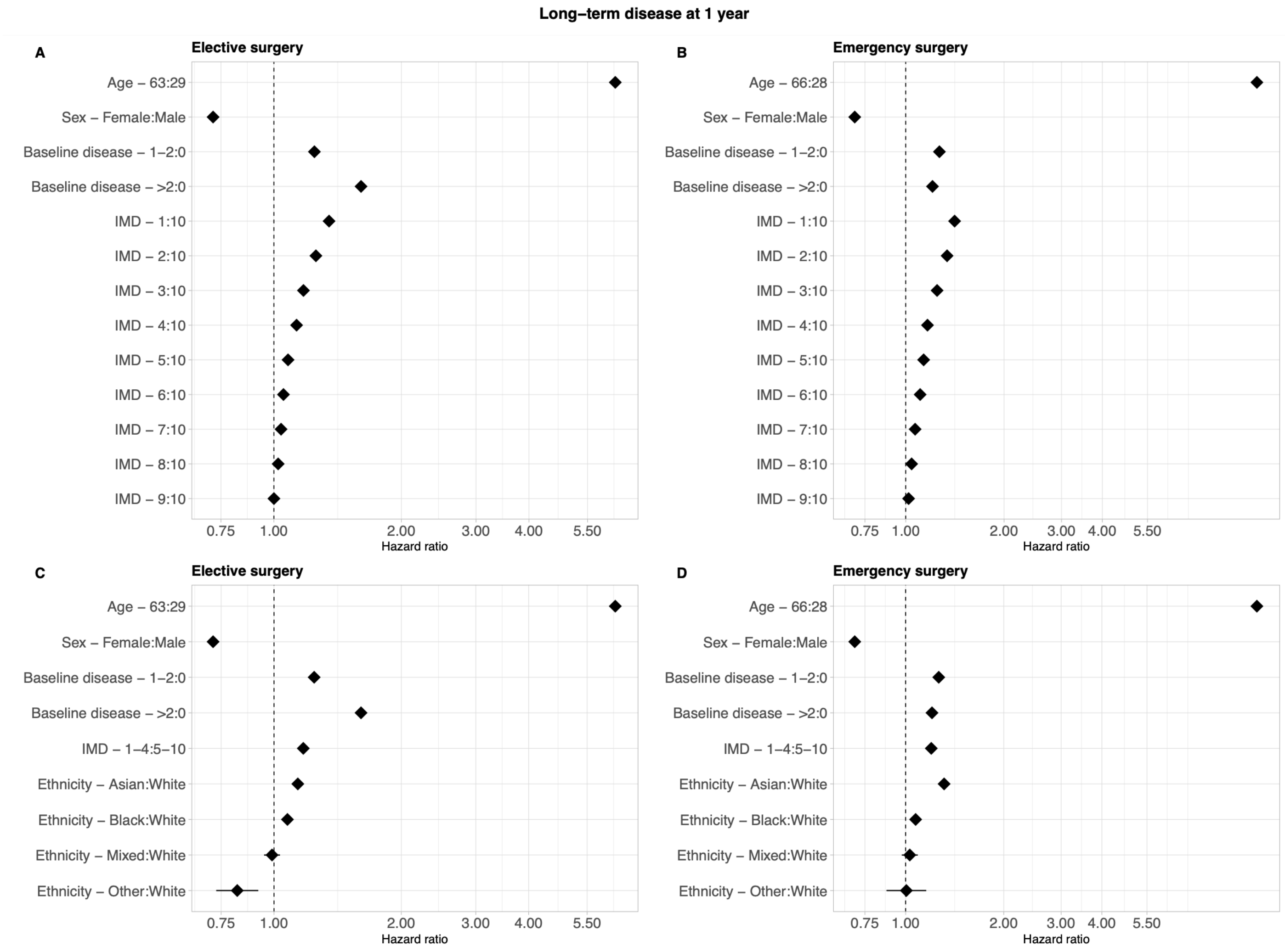

**Figure S6. Forest plots showing adjusted HRs of developing long-term disease (excluding cancer) following surgery at one year. Panel A: Elective surgery by IMD decile, B: Emergency surgery by IMD decile, C: Elective surgery by ethnicity, D: Emergency surgery by ethnicity.** Analysis of Index of Multiple Deprivation (IMD) decile compared to 10 (least deprived) included age, sex (excluding unknown), and baseline number of long-term diseases. Analysis of ethnicity compared to White included age, sex (excluding unknown), baseline number of long-term diseases, and most deprived IMD group (IMD 1-4). Hazard ratio for age comparing 75th to 25th centile. X-axis shown in log scale.

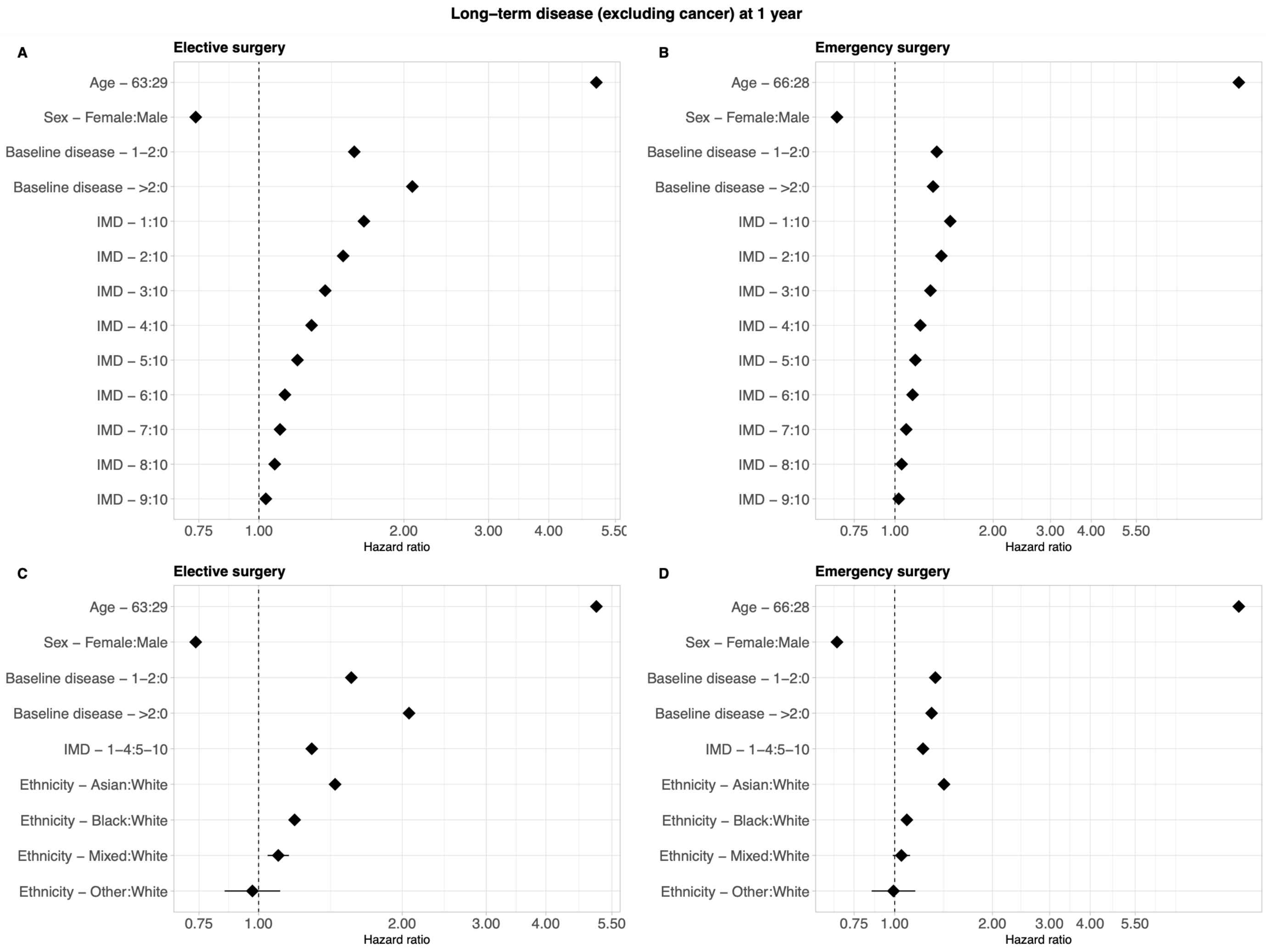

**Figure S7. Forest plots showing adjusted HRs of developing long-term disease (excluding cancer) following surgery at three years. Panel A: Elective surgery by IMD decile, B: Emergency surgery by IMD decile, C: Elective surgery by ethnicity, D: Emergency surgery by ethnicity.** Analysis of Index of Multiple Deprivation (IMD) decile compared to 10 (least deprived) included age, sex (excluding unknown), and baseline number of long-term diseases. Analysis of ethnicity compared to White included age, sex (excluding unknown), baseline number of long-term diseases, and most deprived IMD group (IMD 1-4). Hazard ratio for age comparing 75th to 25th centile. X-axis shown in log scale.

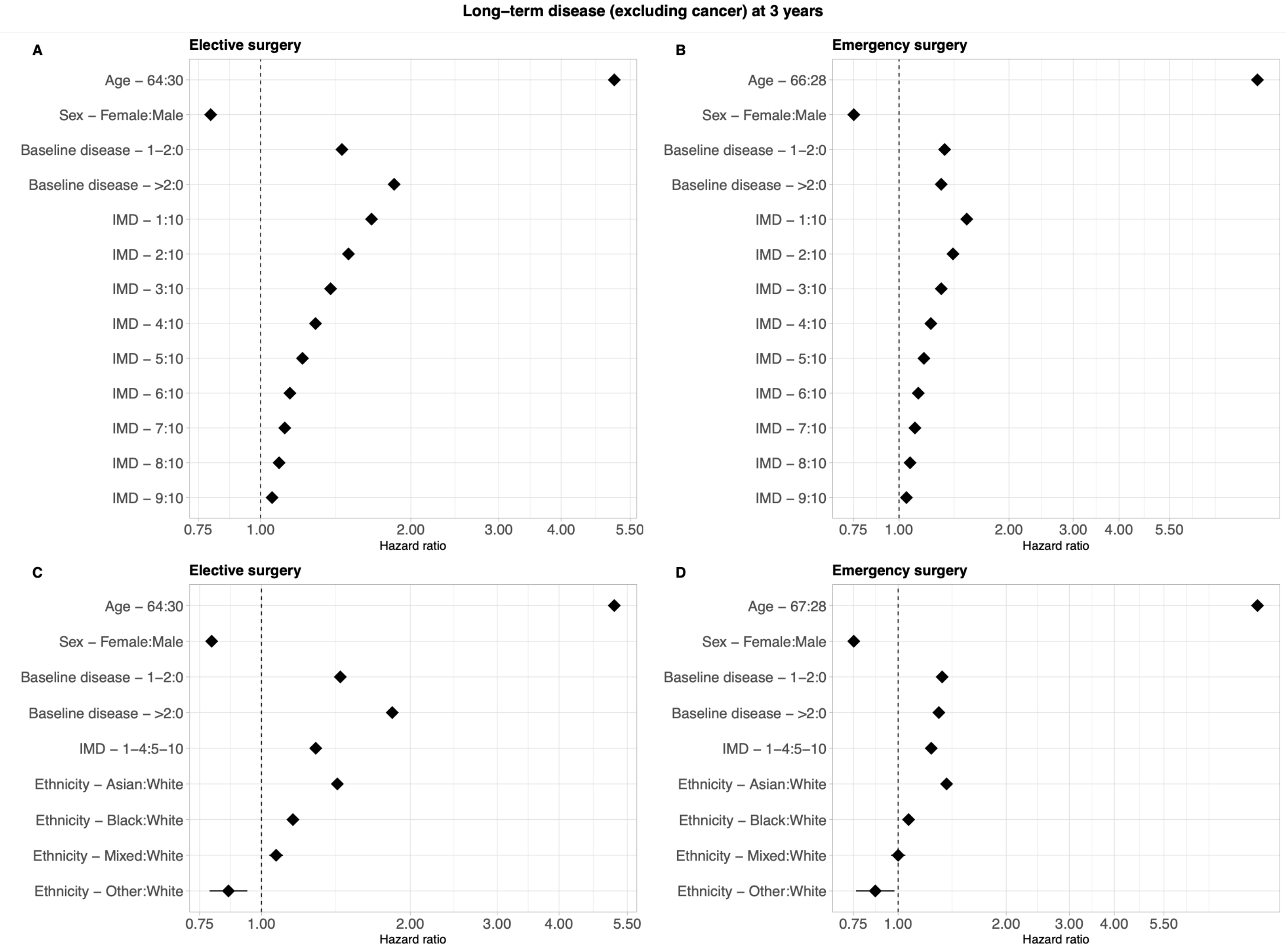

**The RECORD statement – checklist of items, extended from the STROBE statement, that should be reported in observational studies using routinely collected health data.**

| | Item No. | STROBE items | Location in manuscript where items are reported | RECORD items | Location in manuscript where items are reported |
|---|---|---|---|---|---|
| **Title and abstract** | | | | | |
| | 1 | (a) Indicate the study's design with a commonly used term in the title or the abstract (b) Provide in the abstract an informative and balanced summary of what was done and what was found | Main text pages 1-2 | RECORD 1.1: The type of data used should be specified in the title or abstract. When possible, the name of the databases used should be included.<br><br>RECORD 1.2: If applicable, the geographic region and timeframe within which the study took place should be reported in the title or abstract.<br><br>RECORD 1.3: If linkage between databases was conducted for the study, this should be clearly stated in the title or abstract. | Main text pages 1-2<br><br>Main text pages 1-2<br><br>Main text pages 1-2 |
| **Introduction** | | | | | |
| Background rationale | 2 | Explain the scientific background and rationale for the investigation being reported | Main text page 4 | | |
| Objectives | 3 | State specific objectives, including any prespecified hypotheses | Main text page 4 | | |
| **Methods** | | | | | |
| Study Design | 4 | Present key elements of study design early in the paper | Main text page 5 | | |
| Setting | 5 | Describe the setting, locations, and relevant dates, including periods of recruitment, exposure, follow-up, and data collection | Main text page 5 | | |
| Participants | 6 | *(a) Cohort study* - Give the eligibility criteria, and the sources and methods of selection of participants. Describe methods of follow-up | Main text page 5, Figure 1 | RECORD 6.1: The methods of study population selection (such as codes or algorithms used to identify subjects) should be listed in detail. If this is not possible, an explanation should be provided. | Main text page 5 |

| | | | | | |
|---|---|---|---|---|---|
| | | *Case-control study* - Give the eligibility criteria, and the sources and methods of case ascertainment and control selection. Give the rationale for the choice of cases and controls<br>*Cross-sectional study* - Give the eligibility criteria, and the sources and methods of selection of participants<br><br>*(b) Cohort study* - For matched studies, give matching criteria and number of exposed and unexposed<br>*Case-control study* - For matched studies, give matching criteria and the number of controls per case | N/A | RECORD 6.2: Any validation studies of the codes or algorithms used to select the population should be referenced. If validation was conducted for this study and not published elsewhere, detailed methods and results should be provided.<br><br>RECORD 6.3: If the study involved linkage of databases, consider use of a flow diagram or other graphical display to demonstrate the data linkage process, including the number of individuals with linked data at each stage. | N/A<br><br>Main text page 5, Figure S1 |
| Variables | 7 | Clearly define all outcomes, exposures, predictors, potential confounders, and effect modifiers. Give diagnostic criteria, if applicable. | Main text pages 5-6 | RECORD 7.1: A complete list of codes and algorithms used to classify exposures, outcomes, confounders, and effect modifiers should be provided. If these cannot be reported, an explanation should be provided. | Supplement Table 1 |
| Data sources/ measurement | 8 | For each variable of interest, give sources of data and details of methods of assessment (measurement).<br>Describe comparability of assessment methods if there is more than one group | Main text page 5 | | |
| Bias | 9 | Describe any efforts to address potential sources of bias | Main text pages 5-6 | | |
| Study size | 10 | Explain how the study size was arrived at | Main text page 5 | | |
| Quantitative variables | 11 | Explain how quantitative variables were handled in the analyses. If applicable, describe which groupings were chosen, and why | Main text pages 5-6 | | |
| Statistical methods | 12 | (a) Describe all statistical methods, including those used to control for confounding | Main text pages 5-6<br><br>Main text pages 5-6 | | |

| | | | | | |
|---|---|---|---|---|---|
| | | (b) Describe any methods used to examine subgroups and interactions<br>(c) Explain how missing data were addressed<br>(d) *Cohort study* - If applicable, explain how loss to follow-up was addressed<br>*Case-control study* - If applicable, explain how matching of cases and controls was addressed<br>*Cross-sectional study* - If applicable, describe analytical methods taking account of sampling strategy<br>(e) Describe any sensitivity analyses | Main text pages 5-6<br><br>N/A<br><br><br><br>Main text page 6 | | |
| Data access and cleaning methods | | .. | | RECORD 12.1: Authors should describe the extent to which the investigators had access to the database population used to create the study population.<br><br>RECORD 12.2: Authors should provide information on the data cleaning methods used in the study. | Main text page 12<br><br>Main text page 5 |
| Linkage | | .. | | RECORD 12.3: State whether the study included person-level, institutional-level, or other data linkage across two or more databases. The methods of linkage and methods of linkage quality evaluation should be provided. | Main text page 5 |
| **Results** | | | | | |
| Participants | 13 | (a) Report the numbers of individuals at each stage of the study (*e.g.*, numbers potentially eligible, examined for eligibility, confirmed eligible, included in the study, completing follow-up, and analysed)<br>(b) Give reasons for non-participation at each stage. | Main text page 7, Figure S1 | RECORD 13.1: Describe in detail the selection of the persons included in the study (*i.e.,* study population selection) including filtering based on data quality, data availability and linkage. The selection of included persons can be described in the text and/or by means of the study flow diagram. | Main text page 5 |

| | | | |
|---|---|---|---|
| | | (c) Consider use of a flow diagram | |
| Descriptive data | 14 | (a) Give characteristics of study participants (*e.g.*, demographic, clinical, social) and information on exposures and potential confounders<br>(b) Indicate the number of participants with missing data for each variable of interest<br>(c) *Cohort study* - summarise follow-up time (*e.g.*, average and total amount) | Main text page 7, Table 1<br><br>Main text page 7, Tables 1-2, Supplement<br>Main text page 5 |
| Outcome data | 15 | *Cohort study* - Report numbers of outcome events or summary measures over time<br>*Case-control study* - Report numbers in each exposure category, or summary measures of exposure<br>*Cross-sectional study* - Report numbers of outcome events or summary measures | Main text pages 7-8, Table 2, Supplement |
| Main results | 16 | (a) Give unadjusted estimates and, if applicable, confounder-adjusted estimates and their precision (e.g., 95% confidence interval). Make clear which confounders were adjusted for and why they were included<br>(b) Report category boundaries when continuous variables were categorized<br>(c) If relevant, consider translating estimates of relative risk into absolute risk for a meaningful time period | Main text pages 7-8, Supplement<br><br>Main text pages 7-8, Supplement<br><br>N/A |
| Other analyses | 17 | Report other analyses done—e.g., analyses of subgroups and interactions, and sensitivity analyses | Main text page 8 |
| **Discussion** | | | |
| Key results | 18 | Summarise key results with reference to study objectives | Main text page 9 |

| | | | | | |
|---|---|---|---|---|---|
| Limitations | 19 | Discuss limitations of the study, taking into account sources of potential bias or imprecision. Discuss both direction and magnitude of any potential bias | Main text page 10 | RECORD 19.1: Discuss the implications of using data that were not created or collected to answer the specific research question(s). Include discussion of misclassification bias, unmeasured confounding, missing data, and changing eligibility over time, as they pertain to the study being reported. | Main text page 10 |
| Interpretation | 20 | Give a cautious overall interpretation of results considering objectives, limitations, multiplicity of analyses, results from similar studies, and other relevant evidence | Main text pages 9-11 | | |
| Generalisability | 21 | Discuss the generalisability (external validity) of the study results | Main text pages 9-11 | | |
| **Other Information** | | | | | |
| Funding | 22 | Give the source of funding and the role of the funders for the present study and, if applicable, for the original study on which the present article is based | Main text page 6 | | |
| Accessibility of protocol, raw data, and programming code | | | | RECORD 22.1: Authors should provide information on how to access any supplemental information such as the study protocol, raw data, or programming code. | Main text page 12 |

*Reference: Benchimol EI, Smeeth L, Guttmann A, Harron K, Moher D, Petersen I, Sørensen HT, von Elm E, Langan SM, the RECORD Working Committee. The REporting of studies Conducted using Observational Routinely-collected health Data (RECORD) Statement. *PLoS Medicine* 2015; in press.

*Checklist is protected under Creative Commons Attribution (CC BY) license.
